# The Immunogenetic Basis of Idiopathic Bone Marrow Failure Syndromes: A Paradox of Similarity and Self-Presentation

**DOI:** 10.1101/2021.05.28.21258028

**Authors:** Simona Pagliuca, Carmelo Gurnari, Hassan Awada, Ashwin Kishtagari, Sunisa Kongkiatkamon, Laila Terkawi, Misam Zawit, Yihong Guan, Thomas LaFramboise, Babal K. Jha, Bhumika J. Patel, Betty K. Hamilton, Navneet S. Majhail, Sofie Lundgren, Satu Mustjoki, Yogen Saunthararajah, Valeria Visconte, Timothy Chan, Chao-Yie Yang, Tobias L. Lenz, Jaroslaw P. Maciejewski

## Abstract

Idiopathic aplastic anemia (IAA) is a rare autoimmune bone marrow failure disorder initiated by a human leukocyte antigen (HLA)-restricted T-cell response to unknown antigens. Immunogenetic patterns associated with self-antigenic presentation remain unclear. Herein we analyzed the molecular landscape of HLA complexes and T-cell receptor (TCR) repertoires of a large cohort of IAA patients and controls. We show that antigen binding sites of class II HLA molecules in IAA are characterized by a high level of structural homology, only partially explained by specific risk allele profiles, implying reduced binding capabilities compared to controls. Few amino acids within the synapsis HLA-DRB1-antigen-TCR, are identified as strongly associated with IAA phenotype. Those structural patterns may affect TCR repertoires, promoting immunological cross-reactivity and autoimmunity. These findings inform on the immunogenetic risk associated with IAA and on general pathophysiological mechanisms potentially involved in autoimmunity.

**Key points:**

- Class II human leukocyte antigen (HLA) loci in idiopathic bone marrow failure (BMF) syndromes are characterized by low functional divergence and decreased peptide binding capabilities, only partially explained by enrichment in risk alleles.
- A superstructure at the interface with the peptide binding site of DRB1 locus, potentially involved in the presentation of self-antigenic specificities, can be identified in BMF patients.
- This immunogenetic pattern may contribute to decrease T-cell receptor repertoire diversity, expand autoreactive T-cell clones and increase autoimmune propensity in BMF.

## Introduction

Among bone marrow failure syndromes (BMF), idiopathic aplastic anemia (IAA) is a hematopoietic stem cell (HSC) disorder mediated by autoimmune T cells. Despite the progress in our understanding of basic disease mechanisms, the strongest evidence for the immune pathogenesis of this disease stems from the successes associated with immune suppressive therapies (IST).^1^ In addition, clinical observations such as evolution of paroxysmal nocturnal hemoglobinuria (PNH) clones escaping from autoimmune selection pressures, as well as somatic loss of human leukocyte antigen (HLA) alleles due to deletion or mutations, further support the immune nature of this disorder.^2,3,4,5,6^

From a pathophysiological point of view, the *primum movens* of bone marrow destruction is thought to be a class I HLA-restricted process, characterized by cytotoxic T lymphocytes (CTLs) recognition of still unknown HSCs antigens.^7,8, 9,10^ Experimental data demonstrating activation of CTLs with oligoclonal expansion and skewing of CD8^+^ T-cell receptor (TCR) repertoire together with an interferon gamma (IFN-y)-driven FAS-mediated apoptosis of HSCs support this hypothesis.^8,11,12,13,14,15^ Furthermore, the imbalance of CD4^+^ subsets with dysfunctional T helper (Th) 1, Th2 and Th17 responses and consequential impairment of regulatory T cells activities have been demonstrated as additional factors contributing to the aberrant auto-reactivity.^16,17,18,19^ However, because the identity of the eliciting antigens has not been ascertained, the laboratory evidence for T cell-mediated pathogenesis, albeit compelling, remains only indirect. In the context of cellular autoimmune reactions, the pivotal role of HLA molecules in mediating the CD8^+^ and CD4^+^ processes is generally established. Specifically, in IAA a mechanistic involvement of HLA is supported by HLA allele predilection and the modes of immune escape *via* somatic reshuffles of HLA locus (e.g., loss or uniparental disomy of chromosome 6p or somatic mutations in class I alleles). ^2,3,5,20,21, 22,23,24^

While external triggers seem essential, genetic disease susceptibility factors (*e*.*g*., immunogenetic polymorphisms) appear to be operative. Unlike in other autoimmune disorders, the enrichment of some class I alleles in IAA remains limited to ethnicity-restricted series,^2^ whereas the impact of class II loci on disease susceptibility (*e*.*g*., DRB1*15:01) has been historically well documented in multiple populations.^25,26,27,28,29,30,31,32,33,34,35,36,37,38^

Conceptually, the predilection of certain HLA alleles could be explained by their structural “suitability” to present specific immunodominant peptides. However, the relatively small contribution of individual risk alleles in defining the etiologic fraction of IAA, scarcely adapts the structure/function relationships into a disease-specific autoantigenic profile.

Recent studies have shown that HLA evolutionary divergence (HED), a metric capturing the pairwise Grantham distance^39^ between the peptide binding sites of two homologous HLA molecules encoded by an individual’s genotype, correlates with the size of the immunopeptidomic spectrum.^40,41,42^ In this virtue, more structurally divergent alleles may confer a proclivity for more efficient T-cell responses. However, such a postulate, albeit well documented in the context of anti-tumor and anti-infectious immune surveillance,^40,41,43,44^ to date has not been explored in the context of an autoimmune disorder such as IAA. A perhaps simplistic assumption would be that higher allelic divergence may increase the probability of self-antigenic presentation, eliciting autoreactive T-cell responses and explaining the association with autoimmune disease phenotypes.

Herein, we sought to understand the role of HLA functional variability in defining the predisposition and the phenotypic traits of IAA and related disorders. To that end, we performed a large case-control study, in which we analyzed the structural divergence of HLA molecules, its association with risk allele profiles, and the presentation of HSC-related immunopeptidomic specificities. We first performed an allele frequency estimation that helped in the identification and confirmation of the alleles more likely associated with the disease. We then examined the impact of the global genotypic and molecular HLA diversity for both class I and II loci on either disease predisposition or characteristics at diagnosis and clinical outcomes. Finally, we studied how risk and divergent allelic profiles influenced the presentation of HSC immunopeptidome and the TCR repertoire characteristics.

## Results

### Association analysis and risk allele imputation

Out of 430 adult patients with IAA and primary PNH followed at our institution, 263 patients with completed outcomes and precisely asserted diagnosis had DNA for next generation sequencing (NGS)-based HLA typing (**Table 1, Fig.S1**). For comparison, we built two different control cohorts: i) 960 healthy subjects (HC) from a prevalently Caucasian North-American population, and ii) 510 patients with myeloid neoplasms (MN), with known HLA genotypes (see methods and supplementary appendix). For the analysis of HLA associations, the frequency of each allele was evaluated according to a dominant genetic model, assessing the association strength with class assignment.^45^ Among 8 loci, 9 class I and II alleles were identified as differentially distributed between HC and IAA/PNH cohort **(Fig. 1A, Table S1**), with 4 alleles significantly enriched in IAA/PNH patients, according to a dominant genetic model: DRB1*15:01 DQB1*06:02, B*07:02 and DQA1*01:02. Analysis of the additive effect confirmed a strong association with the disease phenotype (**Fig.1B, Table S2**). The distribution of the significant 4 risk alleles did not differ among IAA and hemolytic PNH groups (**Fig.S2A**). To determine whether the presence of each IAA risk allele may influence the course of the disease, we performed a logistic regression univariate analysis, which did not reveal any association with malignant progression to myeloid neoplasia (MN) and/or PNH evolution (**Fig.S2B-D**). However, the presence of DQB1*06:02 showed lower odds in terms of response to IST (OR: 0.48 [95%CI 0.26-0.90], p=0.028, **Fig. S2B-D**). By comparison, none of the risk alleles in study were enriched in patients with MN (**Table S3**). Of note is that those 4 alleles together with A*03:01 and C*07:02 belonged to the ancestral haplotype 7.1,^46^ which was equally distributed across patient and control groups (4% vs. 5%, respectively, p=0.753). Carriers of at least one risk allele were enriched in the IAA vs. HC with 55% of the cases harboring ≥1 risk allele (vs. 39% in HC, OR: 1.94 [95%CI 1.47-2.55], p<0.0001, **Fig.1B**). Furthermore, DRB1*15:01 was associated with DQB1*06:02 in 84% of cases (patients and HC), as a result of the strong linkage disequilibrium. The frequency of homozygous for class II risk alleles in IAA group was higher than in HC, underscoring the role of the “allelic dose” in disease predisposition (**Fig.1C**). Risk allele associations, assessed with binomial regression models, were stronger when considering subgroups of patients with increased autoimmune propensity including i) responders to immunosuppression, ii) age >20yrs.; iii) IAA with PNH clone; iv) all of the above characteristics (**Fig. 1D, Table S4)**.

**Table 1:** Patient characteristics.

| Table 1: Patient characteristics |  |  |
| --- | --- | --- |
| All | N | 263 |
| Age (years) | median (IQR) | 44 (27-62) |
| Race | Caucasian | 228 (86%) |
|  | African American | 23 (8%) |
|  | Asian | 9 (4%) |
|  | Multiracial | 3 (2%) |
| Gender | Female | 132 (50.2%) |
|  | Male | 131 (49.8%) |
| Disease phenotype at diagnosis | IAA +/- non-hemolytic PNH clone | 222 (85%) |
|  | IAA + hemolytic PNH clone | 14 (5%) |
|  | Primary hemolytic PNH | 27 (10%) |
| Severity IAA | Severe | 173 (73%) |
|  | Moderate | 63 (27%) |
| Response to IST* (first line) | CR/PR | 135 (69%) |
|  | NR | 59 (31%) |
| Secondary PNH from IAA | N (%) | 30 (13%) |
| Progression to AML/MDS | N (%) | 32 (12%) |
| Time to secondary PNH | median (IQR) | 49.2 (26.7-122.6) |
| Time to progression AML/MDS | median (IQR) | 39.7 (20.3-71) |
| Follow-up (months) | median (IQR) | 70.8 (30-140) |
| **Abbreviations:** IQR: Interquartile range, IAA: Idiopathic aplastic anemia, PNH: Paroxysmal Nocturnal Hemoglobinuria; AML: Acute Myeloid Leukemia; MDS: Myelodysplastic syndrome; IST: Immunosuppressive treatment * **Overall 194 patients received IST** |  |  |

**Figure 1:**
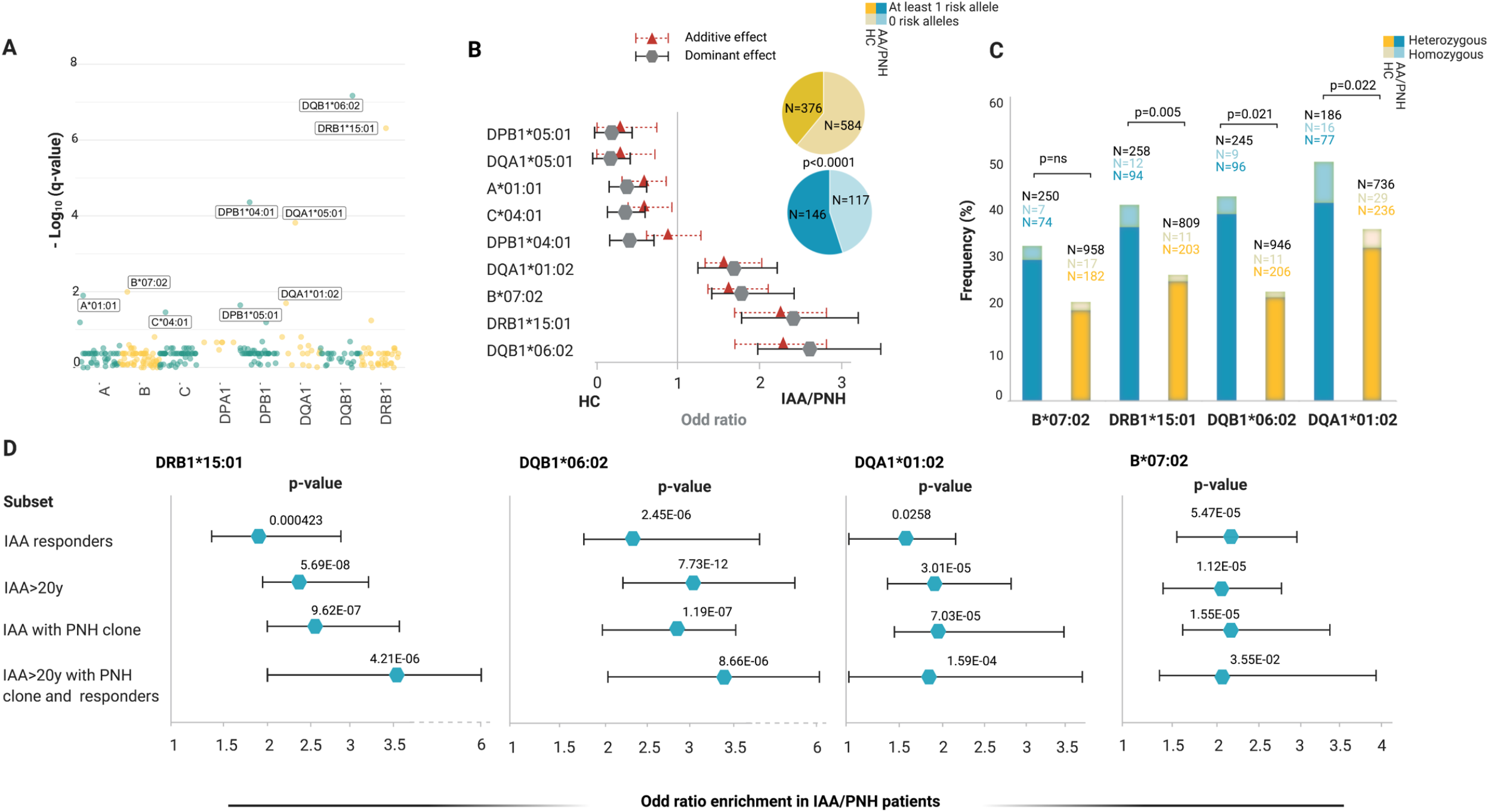
Risk allele profile analysis in idiopathic aplastic anemia and paroxysmal nocturnal hemoglobinuria patients. Abbreviations: HED: HLA Evolutionary divergence; MN: myeloid neoplasia; IAA: Idiopathic aplastic anemia; PNH: paroxysmal nocturnal hemoglobinuria; ns: non-significant. A) Scatterplot representing the negative logarithm base 10 of the adjusted p-values (q-value, Benjamini and Hochberg correction) resulting from the allele association analysis (see methods). Alleles with significantly different genotypic distributions are labeled according to a dominant genetic model. B) Forest plot reporting the odd ratios (OR) defining estimated effect size of alleles enriched in healthy controls (protective) or in patients (risk). Gray markers describe OR resulting from the analysis of genotypic frequencies (dominant model); red triangles depict the OR deriving from analysis of allelic frequencies (additive model). The pie charts illustrate the distributions of subjects with at least 1 risk allele (darkest colors) or without any risk allele (brighter colors). C) Barplot depicting the distribution of heterozygous (darkest) and homozygous (brighter) for the 4 risk alleles in controls and patients. Two-sided Fisher test is applied to test the significance of associations with phenotype. D) Forest plots showing the results of the binomial logistic regression analysis predicting the likelihood of each risk allele association with an aplastic anemia “immune-enriched” phenotype. HC cohort (N=960) was used as comparator group. All (N=263); IAA Responders (N=140); IAA>20 y (N=216); IAA with PNH clones (N=135); Figure 2

When analyzing the allelic frequencies in the pure Caucasian group besides to confirm the strong association with DRB1*15:01 and DQB1*06:02 and the enrichment in B*07:02 and DQA1*01:02 in IAA/PNH patients, we observed a slightly increased frequency of few additional more rare alleles (among which the previously reported B*14:02, see supplementary appendix).^2^

### Low class II HLA divergence as an immunogenetic determinant in IAA patients

We applied HED concept to explore the immunogenetic configuration of IAA and PNH patients, taking into account the risk allele background and computing HED metrics for all classical I and II loci (A, B, C, DRB1, DQB1, DPB1). Based on the previously validated locus-specific HED metrics, for class II divergence computation, we accounted for β-chains only, because of the greater variability of α- chains in terms of peptide binding sites.^41^ Genotypic differences did not impact the class-related homozygosity configurations of the three cohorts, with subjects with at least one homozygous locus per class being equally distributed in the groups (**Fig.2A,B**). When we investigated the organization of global class I and II HED (mean of HED scores for each class) in patients and control groups, no differences were observed for class I (IAA vs. HC: p=0.411 or IAA vs MN p=0.189, **Fig.2C**). Conversely, a lower mean class II HED was found in IAA/PNH cohort vs. HC (p=0.033; **Fig.2D**), involving specifically DRB1 (adj. p=0.028) and DQB1 (adj. p=0.028) loci **(Fig.2E, F**). Associations with lower class II and locus-specific divergence were more evident when we applied generalized linear regression models to predict the risk of immune-enriched disease phenotypes (**Fig.3A, Table S5**). This pattern was confirmed also when considering only Caucasian subjects (see supplementary appendix and Table S6). No differences compared to controls instead were found when analyzing mean class I and class II HED metrics in MN cohort (**Fig. 2A-B**, of note is that MN were characterized by a higher HED in locus B compared to HC).

**Figure 2:**
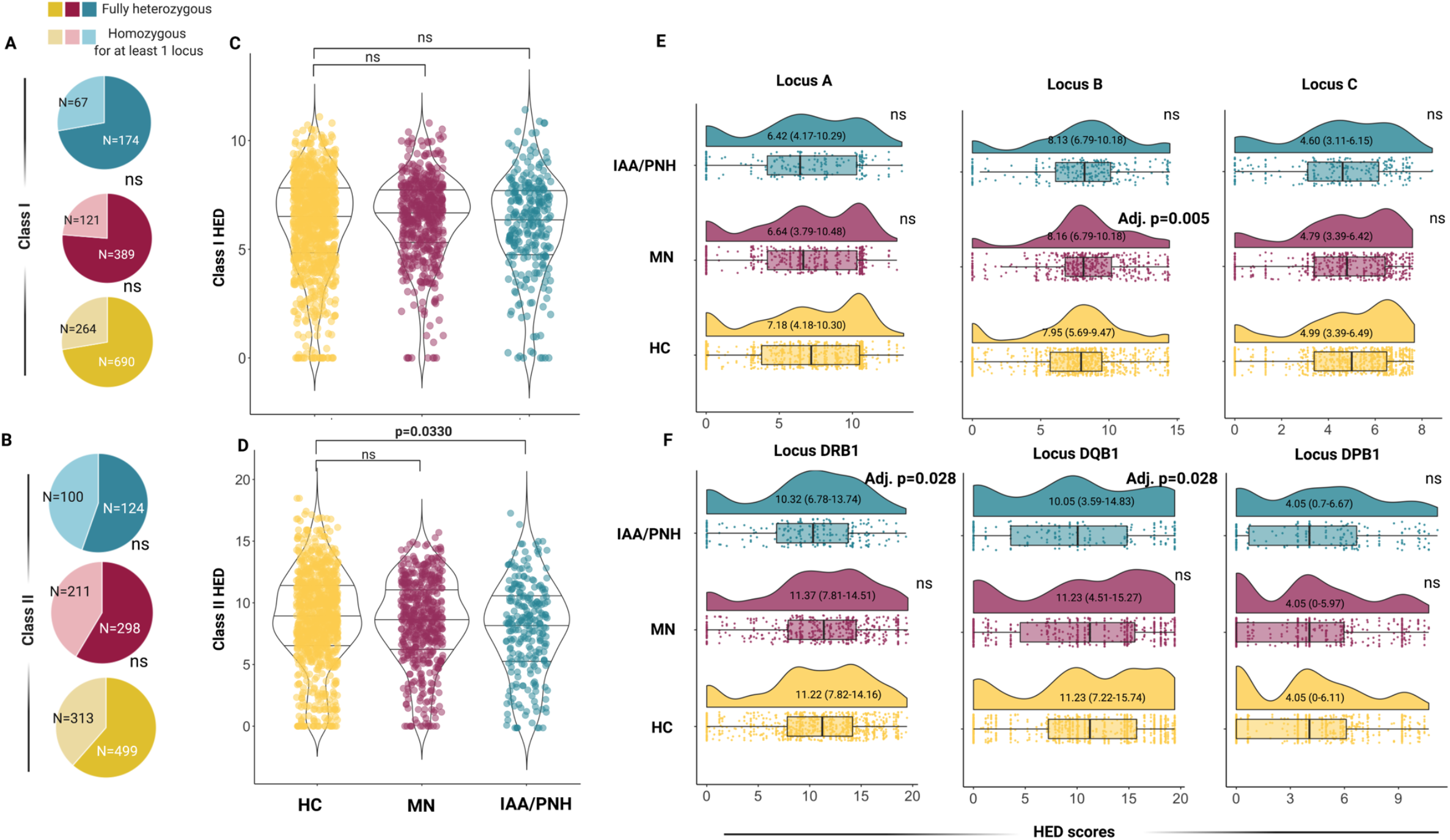
Distribution of evolutionary divergence HLA genotypes in case and control cohorts. Abbreviations: HED: HLA Evolutionary divergence; MN: myeloid neoplasia; IAA: Idiopathic aplastic anemia; PNH: paroxysmal nocturnal hemoglobinuria; ns: non-significant. A) Pie charts representing the distribution of individuals fully heterozygous or homozygous for at least one allele for class I HLA genotypes (yellow: healthy controls; purple: myeloid neoplasia; blue: idiopathic bone marrow failures). Pairwise comparisons with healthy control group; two-tailed Fisher exact test. B) Pie charts representing the distribution of individuals fully heterozygous or homozygous for at least one allele for class IIB HLA genotypes. Pairwise comparisons with HC group; two-tailed Fisher exact test. C) Violin-plot representing the distribution of mean class I HED scores. Each dot denotes the mean value per individual. Wilcoxon signed rank test, comparing each cohort with healthy controls. D) Violin-plot representing the distribution of mean class II HED scores. Each dot denotes the mean value per individual. Wilcoxon signed rank test is used to calculate the p-value. Each cohort is compared with healthy controls. E) Raincloud plots showing the distribution of the HED scores for each class I locus. Each graph is composed by i) a density plot representing the distribution of the metric, ii) a horizontal boxplot showing the median, the interquartile and the overall ranges of HED scores and iii) and a scatter distribution capturing the number of individuals profiled for the given locus and each HED value. Median and IQR are reported in each density plot. Wilcoxon signed rank test is used to calculate the p-value. Multitest-adjustment according to the Benjamini and Hochberg correction is computed and significativity is showed as adjusted p-value. HED scores of each cohort were compared with healthy controls. F) Raincloud plots showing the distribution of the HED scores for each class II locus, as for E.

When the additive effect of the presence of risk alleles on class II HED configurations was analyzed, we observed that lower locus specific HED was independently associated with IAA/PNH phenotype (**Fig.3B**). Interestingly, HC carrying DRB1*15:01 had lower class II and locus-specific HED compared to non-carriers **(Fig.S3A, B)**, whereas in IAA both class II and DRB1 divergences were globally reduced either in presence or in absence of this risk allele (**Fig.S3C**,**D**). Nevertheless, patients without DRB1*15:01 had a significantly lower mean class II (p=0.0005) and DRB1 HED (p<0.0001) compared to HC (**Fig.3C, D**). A similar pattern was seen in DQB1 HED among non-carriers of DQB1*06:02 (**Fig. 3E, F**). When for comparison purposes we examined another autoimmune disease cohort (type 1 diabetes [T1D]), we also found a lower divergence in class II loci (p= 0.00058) and in DRB1 locus compared to HC (p= 0.00104; **Table S6; Fig.S4 A, B, C**). Binomial regression analysis confirmed lower HED in DRB1 locus as a predictor of T1D (OR: 0.97 [95%CI 0.95-0.99], p= 0.00235, **Fig S4D**). Indeed, it is noteworthy the established role of DRB1*15:01 as a protective allele in T1D (**Fig. S4D**), as well as the different risk allele profile dominated by DRB1*03:01 and DRB1*04:01.^47^,^48^,^49^

**Figure 3:**
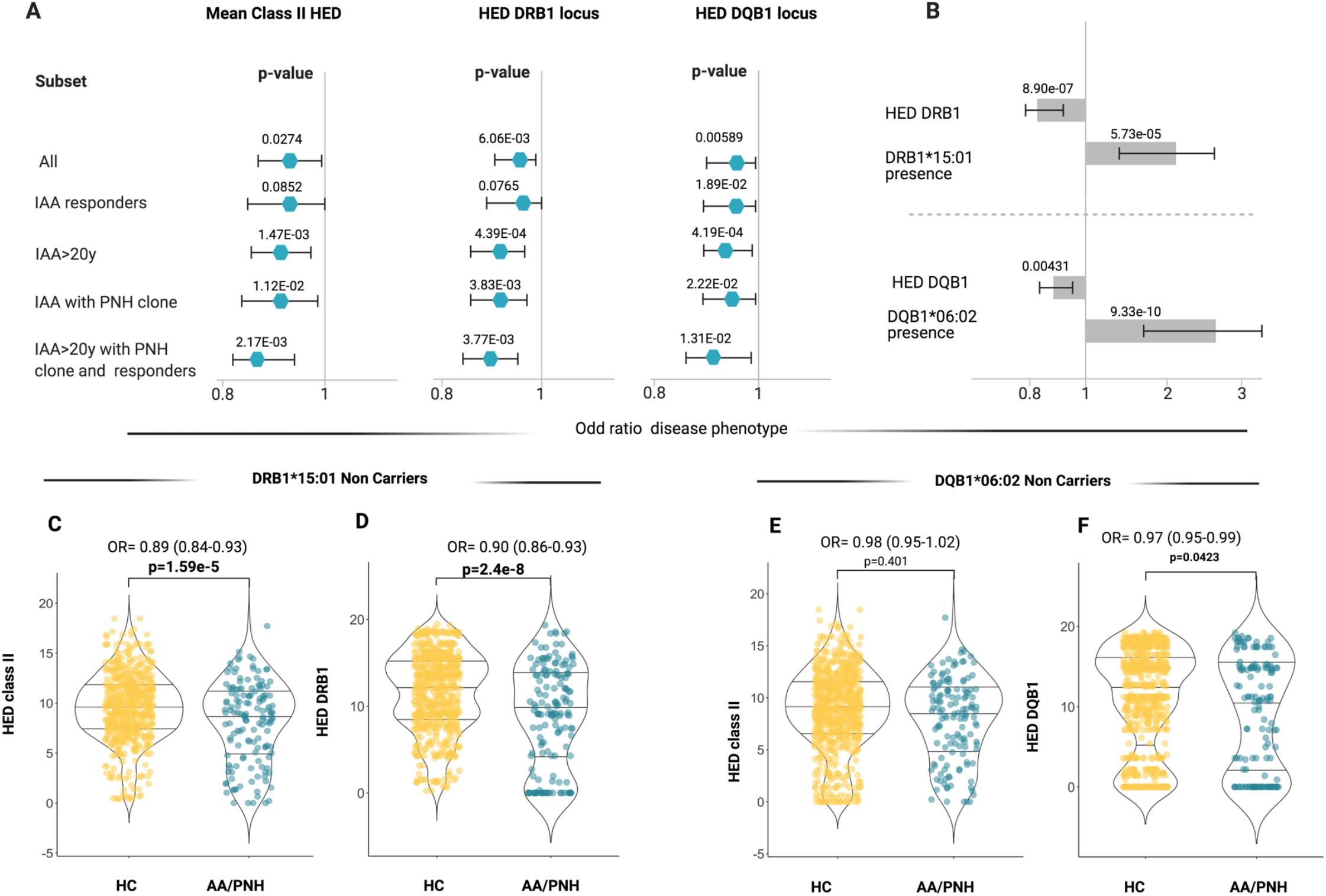
Binomial logistic regression analysis predicting the association between class II HED scores and aplastic anemia phenotypes. Abbreviations: HED: HLA Evolutionary divergence; IAA: Idiopathic aplastic anemia; PNH: paroxysmal nocturnal hemoglobinuria; HC: healthy controls A) Forest plots showing the results of the binomial logistic regression analysis predicting the likelihood of class II and locus specific HED scores of being associated with an aplastic anemia “immune-enriched” phenotype. All (N=263); IAA Responders (N=140); IAA>20 y (N=216); IAA with PNH clones (N=135); IAA>20 y with PNH clone and Responders (N=59). B) Multivariable logistic regression analysis testing the independent effect of HED and risk alleles on idiopathic bone marrow failure phenotype. The length of the bars indicates the odd ratio, the error bars show the 95% confident intervals, the numbers on the bars depict the p-values resulting from the likelihood ratio test. Two distinct models are built for DRB1 and DQB1 locus. C) Violin plots representing the mean class II HED distribution across healthy controls and aplastic anemia patients not carrying DRB1*15:01. Wilcoxon signed rank test was used to calculate the p-value. D) Violin plots representing the DRB1 HED distribution across healthy controls and aplastic anemia patients not carrying DRB1*15:01. Wilcoxon signed rank test was used to calculate the p-value. E) Violin plots representing the mean class II HED distribution across healthy controls and aplastic anemia patients not carrying DQB1*06:02. Wilcoxon signed rank test was used to calculate the p-value. F) Violin plots representing the DQB1 HED distribution across healthy controls and aplastic anemia patients not carrying DQB1*06:02. Wilcoxon signed rank test was used to calculate the p-value.

To identify alleles structurally similar to those identified as risk alleles in class II, we simulated the range of HED scores between DRB1*15:01/DQB1*06:02 and the pool of alleles present in DRB1 and DQB1 loci in IAA/PNH patients and HC (**Fig.4A-C**). In DRB1 locus, a lower divergence with DRB1*15:01 was obtained for alleles within DRB1*15, DRB1*16, DRB1*04 and DRB1*01 supertypes (**Fig.4A; Table S7**). Combined genotypic frequencies of those DRB1*15:01-like alleles were higher in IAA/PNH population (OR: 1.89 [95%CI: 1.38-2.61], p=7.22e-05) and in cases with immune-related phenotypes compared to HC (**Fig.4B**). Analogous results were observed for DQB1 locus and DQB1*06 supertype (**Fig.4C, D; Table S8**).

**Figure 4:**
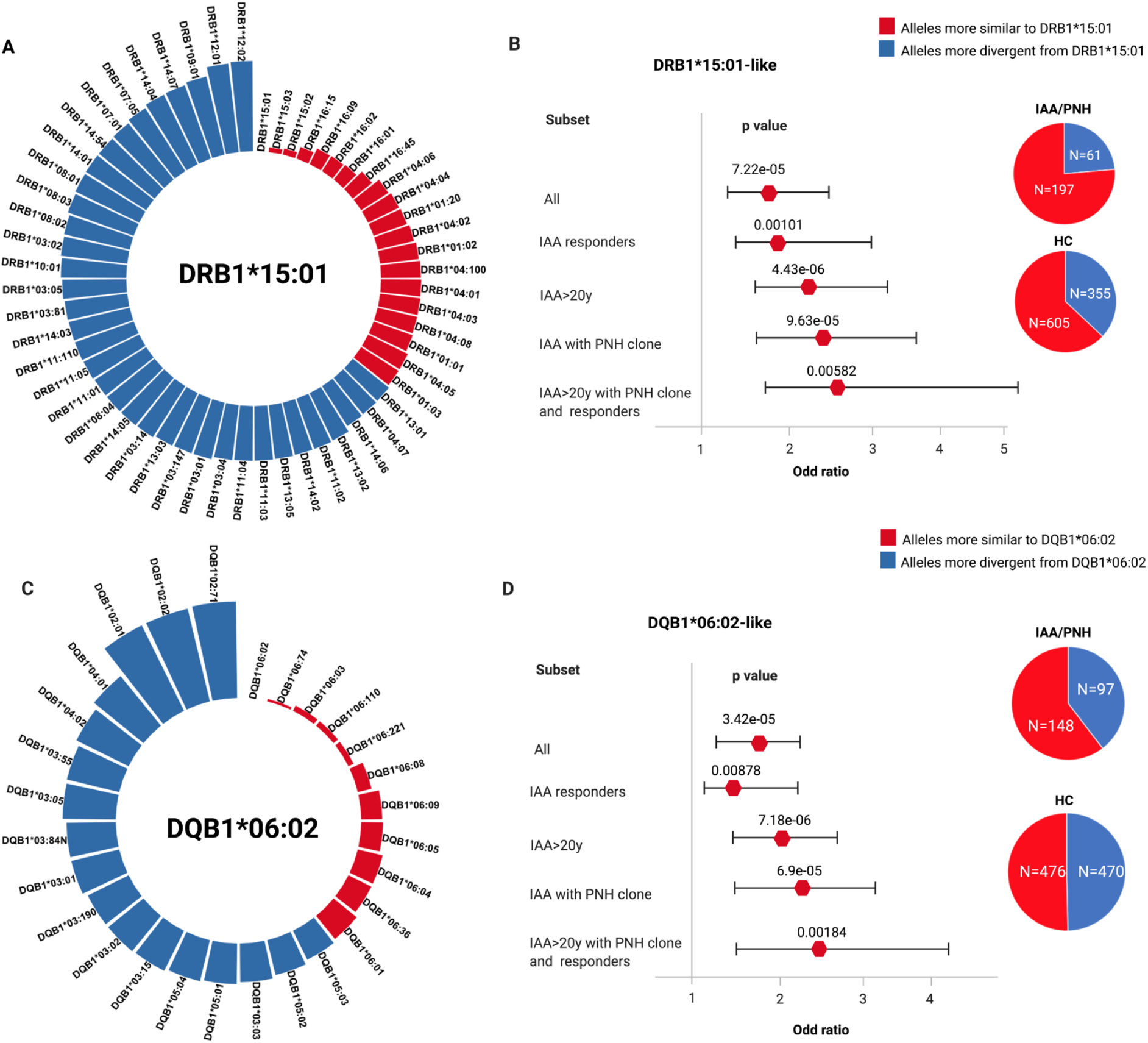
Simulated structural divergence between each class II risk allele and the pool of alleles present in DRB1 and DQB1 loci in patients and controls. Abbreviations: HED: HLA Evolutionary divergence; IAA: Idiopathic aplastic anemia; PNH: paroxysmal nocturnal hemoglobinuria; HC: healthy controls A) Circle graph representing the simulated divergences between DRB1*15:01 and each allele present in DRB1 locus of IAA/PNH patients and HC. Red bars illustrate the alleles more similar to DRB1*15:01 (divergent pairs located under the 25th percentile cutoff of the simulated distribution). B) Forest plots showing the results of the binomial logistic regression analysis predicting the association between the presence of DRB1*15:01-like alleles and aplastic anemia “immune-enriched” phenotypes. HCs (N=960) were used as comparator group. Pie charts describe the distribution of alleles more similar and more divergent from DRB1*15:01 in patients and controls. All (N=263); IAA Responders (N=140); IAA>20 y (N=216); IAA with PNH clones (N=135); IAA>20 y with PNH clone and Responders (N=59). C) Circle graph representing the simulated divergences between DQB1*06:02 and each allele present in DQB1 locus of IAA/PNH patients and HC. Red bars illustrate the alleles more similar to DQB1*06:02 (divergent pairs located under the 25th percentile cutoff of the simulated distribution). IAA>20 y (N=216); IAA with PNH clones (N=135); IAA>20 y with PNH clone and Responders (N=59). D) Forest plots showing the results of the binomial logistic regression analysis predicting the association between the presence of DQB1*06:02-like alleles and an aplastic anemia “immune-enriched” phenotype. HCs (N=960) were used as comparator group. Pie charts describe the distribution of alleles more similar and more divergent from DQB1*06:02 in patients and controls.

When we studied whether HLA functional divergence may influence characteristics and outcomes of IAA/PNH patients, we found that class II HED scores correlated directly with the size of PNH clone at diagnosis (**Fig.S5A-B;** p= 0.00286, r^2^=0.036) and indirectly with age at disease onset (**Fig.S5C**-**D;** p=0.0013, r^2^=0.021). Univariable cox regression models (based on binomial categorization of mean class I and II HED according to the 50^th^ percentile in HC, **Fig.S5E-K**), demonstrated lower probability of survival (p=0.011), higher risk of progression to MDS/AML (p=0.043) and a lower probability of PNH evolution (p=0.0004, **Fig.S5J**,**L**) in patients with lower mean class II HED, while no impact was seen for class I HED (**Fig.S5G**,**I**,**J**).

### Recursive analysis of the antigen binding site in DRB1 and DQB1 loci

To investigate whether low divergent patterns could rely on a specific amino acid composition of the peptide binding site of class II HLA molecules, we analyzed the amino acid structure in the antigen binding site (encoded by exon 2) of DRB1 and DQB1 loci. In DRB1 peptide binding groove, 30 out of 89 amino acid positions were variable. By applying a recursive approach we found strongly correlated with the IAA/PNH phenotype 7 amino acids (**Fig. 5A**,**B; Table S9**), enriched in DRB1*15 group and in all the alleles structurally similar to this supertype. Of note is that most of those residues were non-polar, possibly affecting the physicochemical configuration of the antigen-binding site of DRB1*15-like complexes. When the same analysis was performed on DQB1 locus (**Fig.S6A**,**B; Table S10**), the majority of variable amino acids in exon 2 were enriched in HC, with the exception of 2 residues of phenylalanine (Phe/F), both belonging to the DQB1*06 supertype and significantly overrepresented in IAA (**Fig.S6A**,**B**).

**Figure 5:**
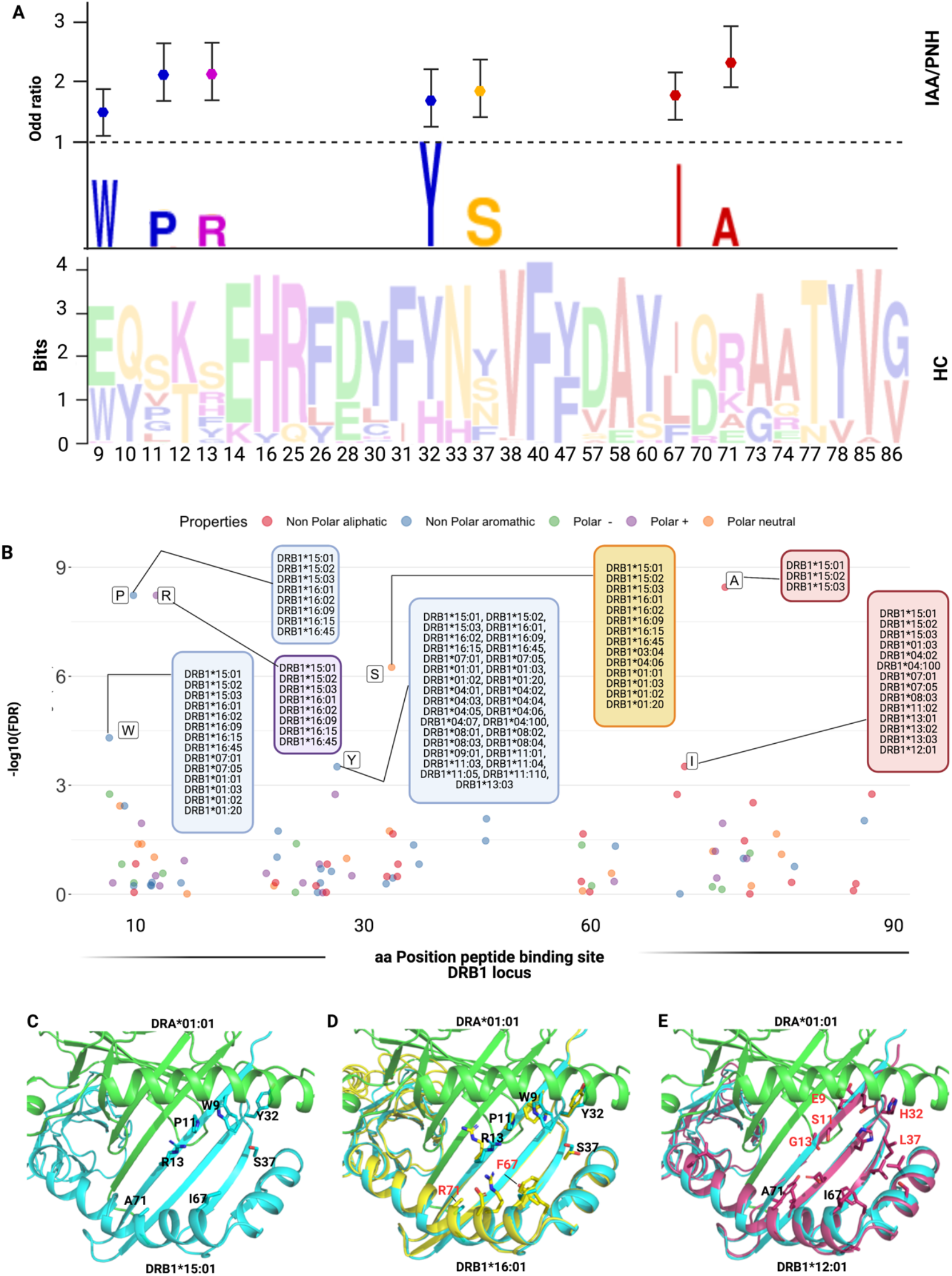
Recursive analysis of the amino acid sequence within the peptide binding site of DRB1 locus. Abbreviations: IAA: Idiopathic aplastic anemia; PNH: paroxysmal nocturnal hemoglobinuria; HC: healthy controls; aa: amino acid A) Lower panel: WebLogo visualization representing the contribution of single aminoacids within the variable portion of the peptide binding site of DRB1 locus. The x-axis indicates each variable position (as per IPD-IMGT-HLA reference). Letters represent each possible amino acid at each given position; Letters’ height illustrates the frequency of each amino acid in healthy control population. Colors indicate the chemico-physical properties as per legend in B. B) Upper panel: stylized visualization of amino acids differentially distributed between HC and IAA/PNH cohorts. Letters’ height illustrates the frequency of each amino acid in disease population. Markers indicate the odd ratios resulting from the logistic regression analysis studying each aminoacidic contribution in determining the phenotype (see methods and Table S7). C) Scatter plot showing the significance level of each variable amino acid in the peptide binding site of DRB1 locus found enriched in IAA/PNH population compared to HC. Each dot represents the negative logarithm base 10 for the adjusted p-value (q-value) referring to each amino acid. The position on x-axis indicate the position within the peptide binding site according to IPD-IMGT-HLA reference). Only the amino acids presenting a q-value<10-e4 are considered significant for this analysis and labelled in the figure. Alleles presenting the indicated amino acid at the given position are indicated in the boxes. Colors represent the chemico-physical properties as per legend. D) Crystallographic structure showing the position of the 7 amino acids significantly enriched in the peptide binding groove of DR molecules of IAA/PNH patients. This 3D structure has been visualized with the PyMOL program based on the structure of DRB1*15:01-DRA*01:01-myelin binding protein (PDB:1BX2) and HLA sequences retrieved from IPD-IMGT/HLA database v. 3.40. Only the 7 amino acids identified in the previous analysis have been highlighted. E) Binding site of DRB1*16:01-DRA*01:01 based on the homology model of DRB1*16:01. The structure of DRB1*16:01 has been superimposed to DRB1*15:01. Residues differing from the risk pattern seen in DRB1*15:01 are colored in red. F) Binding site of DRB1*12:01-DRA*01:01 based on the homology model of DRB1*12:01. The structure of DRB1*12:01 has been superimposed to the structure of DRB1*15:01. Residues differing from the risk pattern seen in DRB1*15:01 are colored in red.

Because of this lower contribution of DQB1-binding site variability in conferring disease phenotype, our next analysis was focused on modeling antigen interactions within DRB1 locus. When we analyzed the crystallographic structure of the complex DRB1*15:01/DRA*01:01 (PDB: 1BX2), the identified amino acids clustered within the right part of the antigen binding pocket (**Fig.5C**). Only two of 7 amino acids were different in DRB1*16:01 (indicating higher structural similarity to DRB1*15:01; **Fig.5D**), while 5/7 residues differed in DRB1*12:01 (more structurally divergent from DRB1*15:01; **Fig.5E**). We also aligned the crystal structures of DRB1*15:01/DRA*01:01 with three peptides known to have affinity for DRB1 molecules (EBV DNA polymerase, vimentin and myelin binding protein [MBP], see methods) and found two different patterns of interaction within the binding groove: while the peptide portions binding in the left part of the groove tended to assume the same backbone conformation (underlying conservation of the physicochemical characteristics among the structures at this interface), peptide portions allocated within the right side of HLA groove assumed more variable conformations (**Fig.6A**). To investigate how this structural configuration could affect the TCR binding, we then constructed a model structure of TCR α and β chains, DRA*01:01-DRB1*15:01 and CD4 on the reported ternary crystal structure of HLA-peptide-TCR-CD4 (PDB: 3T0E).^50^ Based on the alignment of these three antigenic models, we found that the variable peptide segments (right portion) interacted mainly with the TCR Vβ chain (**Fig.6B, C**). These findings provided a proof-of-concept for the importance of this consensus structure in accommodating the interactions with antigen and T-cell specificities within DRB1 locus.

**Figure 6:**
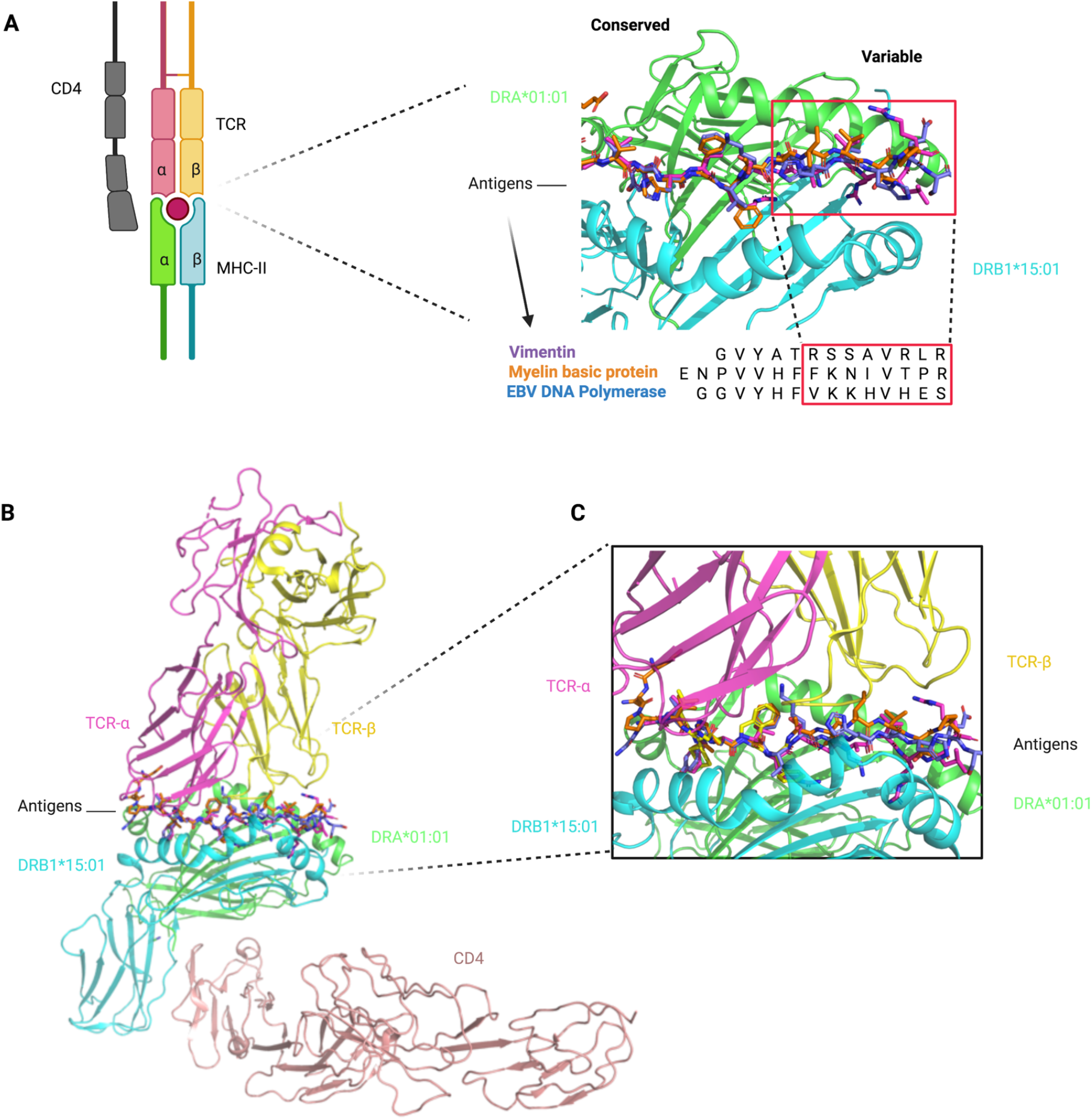
Structural insight into DRB1-antigen-TCR interactions. Abbreviations: TCR: T-cell receptor; MHC: major histocompatibility complex, MBP: myelin binding protein; EBV: Epstein barr virus A) Peptides of vimentin (UniProt: P08670, VIME_HUMAN, 59-71), EBV DNA polymerase (UniProt: P03198, DPOL_EBVB9, 628-641), MBP (UniProt: P02686, MBP_HUMAN, 217-231) at the HLA binding site based on the alignment of crystal structures of DRB1*15:01-DRA*01:01/MBP (217-231) (PDB: 1BX2), DRB1*14:02-DRA*01:01/ vimentin (amino acid positions: 59-71, PDB:1H15) and DRB5*01:01-DRA*01:01/EBV DNA polymerase (628-641) (PDB:6ATF). The red squares indicate the peptide portions presenting with more conformational variability (in interaction with the right site of the HLA binding groove). This 3D structure has been prepared with PyMOL using the crystal structure of DRB1*15:01-DRA*01:01/MBP (PDB: 1BX2). B) Modeled ternary structure of HLA-Antigen-TCR-CD4. C) Detail of the interaction interface in HLA-antigen-TCR. The three antigenic structures are aligned as shown above. The risk amino acid pattern within the right side of the binding groove interacts with a more variable antigenic portion that contacts directly with the TCR beta chain (software PyMOL).

### Quantitative thresholds for self-antigenic presentation

To explore the binding capacity of self-generated peptides possibly involved in AA pathogenesis, we built an *in silico* HSC-specific proteomic reference and we analyzed all the different DRB1 and DQB1 molecules present in IAA and HC cohorts. We generated 15-mer peptides from 40,614 transcripts assembled from 7724 previously identified HSC proteins.^51^ For each sequence, the mean number of strong and weak binders (see methods) was determined across all the DRB1 alleles, along with their proteogenomic spectrum (**Table S11**). Overall, about 10% of this proteomic reference was found capable of generating self-peptides suitable for the binding of DRB1 or DQB1 molecules (all peptides with a percentile rank of eluted ligand prediction score <5% -corresponding to strong and weak binders-were considered for this analysis, **Fig.7A, B**). HLA molecules belonging to the same locus had similar proteogenomic spectra of derived binders. However, quantitative differences in the number of predicted binders were observed across all the alleles (**Table S11 & S12**). In particular DRB1*15:01 along with other alleles was identified as structurally similar to the DRB1*15 supertype and was characterized by lower binding capacities compared to other alleles (**Fig.7B, C**). Analysis of the distribution of the number of binders in HC and IAA/PNH groups showed significantly lower binding capacities in DRB1 locus in patients vs. controls (**Fig.7D**). This pattern was observed also in the subgroup of homozygous individuals (HED score 0; **Fig.7E**). For DQB1 locus immune-peptidomic analysis we accounted for the genotypic associations with DQA1 locus and thus we considered only individuals with known DQA1 allele. DQB1*0602-DQA1*01:02 was predicted as one of the haplotypes with the lowest binding capacities (**Fig. S7A-D**) and its propensity to bind HSC self-peptides was decreased compared to other DQB1*06:02/DQA1 combinations (**Fig.S7B**).

**Figure 7:**
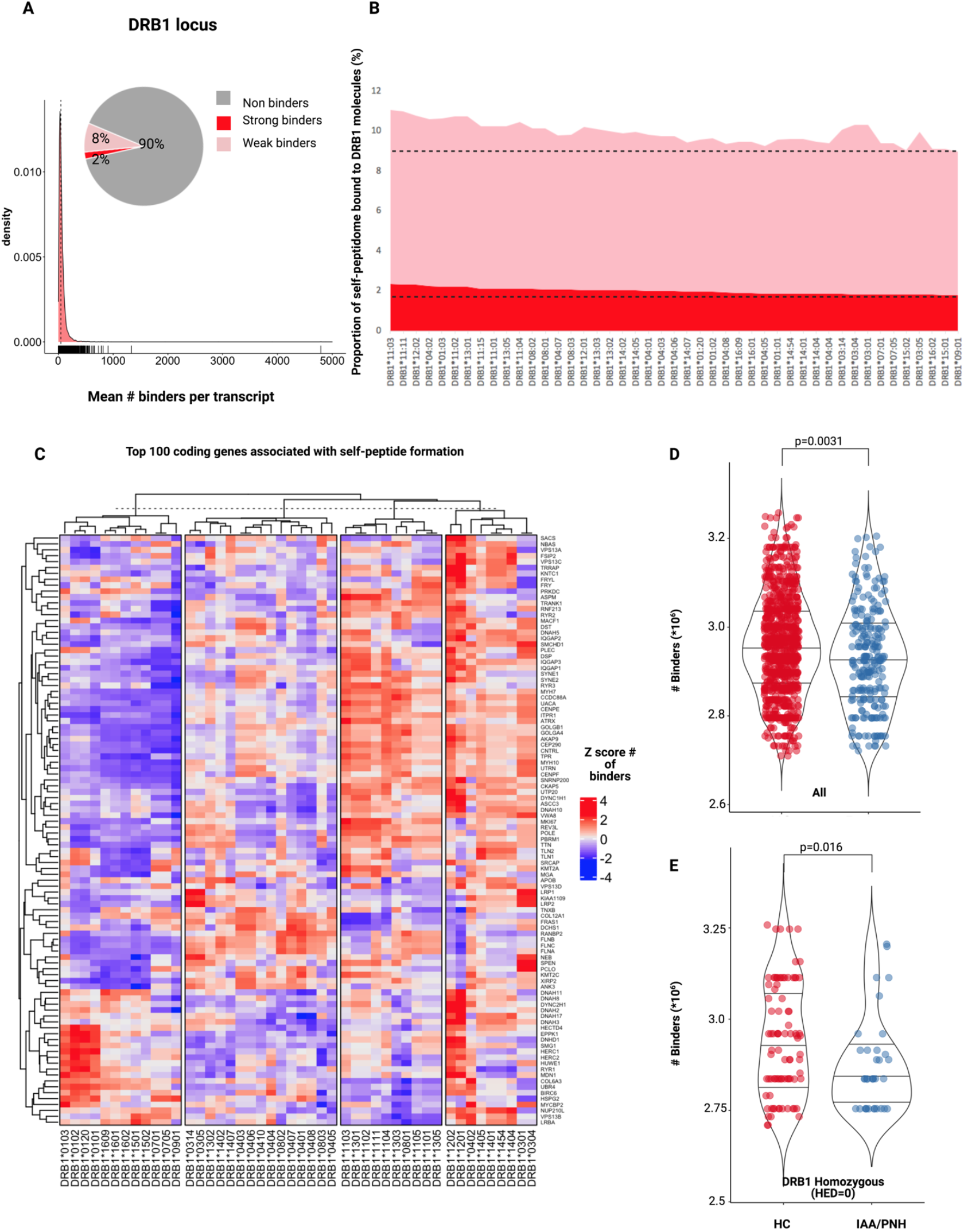
Hematopoietic stem cell specific immune-peptidome binding capacities of the DRB1 molecules. Abbreviations: IAA: Idiopathic aplastic anemia; PNH: paroxysmal nocturnal hemoglobinuria; HC: healthy controls; HSC: hematopoietic stem cell; HED: HLA evolutionary divergence. A) Density plot showing the mean number of binders per each transcript (average of number of strong and weak binders for all DRB1 molecules in study generated from the HSC-specific proteomic reference). The pie chart indicate the proportion of binders and non-binders for the whole reference. Overall 10% of this proteomic repertoire is able to generate potential self-peptides. B) Filled 2D area plot representing the binding capacities of each DRB1 molecule in study for the self peptidomic HSC reference. Y-axis indicate the percentage (%) of self-peptides predicted to bind DRB1 molecules. Red portion indicate strong binders, pink portion of the graph depicts instead weak binders (see methods). C) Heatmap showing the top-100 coding genes associated with the self-peptidome formation. Each cell represents the Z-score of the number of binders predicted of each transcript for a given DRB1 molecule. D) Distribution of the number of binders in HC and IAA/PNH cohorts. Wilcoxon signed rank test. E) Distribution of the number of binders in HC and IAA/PNH homozygous subjects (HED=0). Wilcoxon signed rank test.

To determine whether the findings described above were reproducible across different ethnic groups and populations with unique HLA distributions, we investigated the genotypic and proteogenomic patterns of the DRB1 locus in of IAA patients (N=37, among which N=30 genotyped at 4-digit level) and healthy controls (N=128) from a Finnish cohort. Consistently with our results, DRB1*15:01 was significantly associated with BMF phenotype (**Fig.S8A**,**B**; OR:5.18; p=0.0001) and divergence in DRB1 locus was lower for IAA patients compared to controls (this configuration was found both for the whole cohort and for non-carriers of DRB1*15:01, **Fig.S8C**,**D**). Importantly, as for the main cohort, also in the Finnish one, DRB1 allele distribution was characterized by lower binding capacities in patients vs. corresponding HC.

### Insights in TCR repertoires

To investigate how risk allele profiles and HED configurations may dictate the patterns of T-cell responses, we performed deep TCR Vβ complementary determining region (CDR3) sequencing for 25 patients with IAA. Patients’ Vβ CDR3 spectra were compared to those of 130 HC. After a down-sampling procedure (see Methods), TCR diversity metrics were calculated. As expected, TCR repertoires were characterized by lower diversity compared to HC (IAA vs. HC: p=5.4e-08; number of unique clonotypes: p=0.012; mean size clonal expansion: p=0.512; **Fig.S9A-C**). Specifically, diversity was lower in both DRB1*15:01 carriers and non-carriers compared to controls (**Fig. S9D-E**). No correlation was found between Vβ TCR diversity metrics and class I or II mean HED (**Fig.S9 F-J**). To evaluate how HED could impact on the autoreactive disease-associated spectra, we first built a comprehensive compendium of all CDR3 sequences with known specificity identified in literature (see Methods) and blasted the CDR3 sequences identified in our cohorts against this dictionary. Overall, within the identified clonotypic portion of the repertoire (<2% of the total), the mean proportion of identifiable autoreactive clonotypes was 14% in IAA vs. 6% in HC (**Fig.S9L**)., while their mean frequency was respectively 0.016% and 0.001% (**Fig.S9M;** p<2e-16). In IAA patients, but not in HC, the productive frequency of those clones inversely correlated with mean class II HED (p=1.332E-19; r^2^= 0.041; **Fig.S9N**), underscoring the clonal expansion of those specificities in patients with lower class II HLA divergence. Almost all of the autoimmunity-associated clonotypic groups were found hyperexpanded in IAA compared to HC (**Fig.S90**).

## Discussion

In IAA, the autoimmune destruction of hematopoietic progenitor and stem cells is an HLA class I-and II-restricted T cell-mediated process. Here, with a comprehensive immuno-proteogenomic approach, encompassing deep NGS of HLA region, TCR sequencing and HSC specific immunopeptidome binding analysis, we intensively assessed HLA structures involved in disease susceptibility and potentially associated with autoimmune propensity. To that end, we used not only a comparative population of HC but also two large disease-control datasets.

The quantitative concept of HED relies on divergent allele advantage, stipulating that structural heterogeneity of HLA alleles allows for a wider spectrum of peptides to be presented and thus a higher probability to mount efficient anti-tumor and anti-infectious responses^.52,53,54,40^ Accordingly, a similar principle should apply to autoimmune diseases, with a higher HED reflecting an increased propensity to T cell-mediated autoimmunity. However, we did not found support for this hypothesis and instead observed that HLA molecules in IAA patients were characterized by a high structural similarity, especially in class II, in part due to enrichment for risk alleles and/or alleles structurally similar to risk alleles. These associations were particularly strong for DRB1 and DQB1 loci. Consistently with previous studies, DRB1*15:01 together with DQB1*06:02 were identified as alleles enriched in IAA and PNH.^26,55,27^ A special mention deserves the fact that HED metrics were found to be low, independently of the presence of risk alleles in IAA/PNH setting. This pattern is explained by the global low divergence of class II HLA molecules seen in BMF cohort. Indeed, in IAA patients the non-risk alleles were more structurally similar to each other and to the risk alleles and thus may have analogous (albeit not completely overlapping) peptide recognition spectra. This may contribute to decrease the diversity and increase the clonal expansion of TCR specificities in IAA repertoire. Further, a lower locus specific HED was independently associated with disease phenotype also when performing generalized linear regression models tracking the additive effect of class II HLA risk alleles. Reinforcing the idea that this pattern may be present across different ethnicities and ancestry groups, we showed reproducibility of such findings in an independent cohort of Finnish IAA/PNH patients. Also, we found the same low HLA divergence pattern in an autoimmune disease characterized by different risk allele associations and in which, of note, DRB1*15:01 is a protective allele.^47^

Since our genetic models were built on a mixed population and did not allow to confirm the class I risk alleles previously identified in a prevalently pediatric Caucasian cohort,^2^ we performed also a race/ethnicity stratified subanalysis. Hence, among alleles formerly recognized, we could show only a slightly significant enrichment in B*14:02 in Caucasian patients from the North American cohort, and again confirmed the strong association with the class II alleles identified in the main analysis, as well as the low divergence characterizing class II loci. It is possible that the above dissimilarities with the previously reported class I associations^2^ rely on the different age composition of our study cohort, prevalently composed by adult patients (see Supplementary considerations).

In order to deeper analyze the superstructures involved in the decreased divergence within the antigen binding sites, we studied the amino acid specificities found in the variable portion of the antigenic groove of DRB1 and DQB1 molecules with a recursive approach. This analysis enabled the identification of few residues significantly associated with IAA/PNH phenotype, mainly located in pockets involved in antigen and TCR β interactions. This analysis, by modeling analogous antigens known to be binders of DRB*15:01 and alleles with structural similarity, allowed us to identify antigen components involved in the preferential self-presentation of BMF patient, potentially associated with impaired T-cell activation.

Consistent with the lower divergence seen in IAA/PNH population, when we analyzed DRB1 and DQB1 binding predictions covering the HSC specific immunopeptidome, we found that for both loci, HLA genotypes had lower binding capacities in patients compared to healthy individuals, both in homozygous and heterozygous settings. If per divergent allele advantage immunocompetence is supposed to be enhanced in case of higher HLA divergence, it is plausible that less divergent loci and, per extension, HLA molecules with lower binding capacities (such as DRB1*15:01 and DQB1*06:02-DQA1*01:02) may increase the risk of immunological cross-reactivity and molecular mimicry with possible pathogen-associated antigens, by that triggering autoimmune diseases. As an example of autoimmune disorder with known self-antigenic specificities, in multiple sclerosis the link among DRB1*15:01, EBV and central nervous system antigens’ molecular mimicry is well established, with indirect evidence that impaired CD4+ presentation may elicit aberrant CD8+ responses and auto-antibody production.^56,57,58,59^

If risk allele profiles did not impact on clinical outcomes, low divergence in class II was associated with an increased probability of malignant progression in IAA/PNH patients. This finding is in line with the idea that anti-tumor surveillance (previously shown as more efficient in highly divergent class I allele pairs)^40^ could encompass also HLA class II-restricted T-cell responses.^60,61,62^ In this setting, less divergent class II β chains may reduce the neoantigen presentation capabilities configuring an immune escape scenario.

Our data demonstrate that in BMFs and potentially in other autoimmune disorders, HLA allele configurations with more specific structural patterns and globally lower functional divergence may contribute to decrease the binding capabilities especially in class II alleles, potentially enhancing antigenic cross-reactivity and, hence, autoimmune propensity.

Taken together our results represent an important advancement in the field of immunogenetics of BMFs with crucial implications also in other autoimmune disorders. Analysis of HLA divergence and identification of DRB1 superstructure may be easily translated in clinical practice for better diagnostic and prognostic orientations paving the way for new therapeutical approaches potentially able to modulate the self-antigenic binding capabilities of class II HLA molecules.

## Methods

### Study design and cohort assemblies

This is a retrospective study evaluating the immunogenomic configuration of antigen-presenting HLA complexes and the TCR repertoires of patients with IAA and PNH (**Fig.S1**). Two hundred sixty-three patients diagnosed with BMF, along with 20 HC were enrolled, and their peripheral blood and marrow specimens were previously stored, based on their consent to participate to institutional translational research protocols. This study was conducted under the institutional review board of Cleveland Clinic (IRB #5024). Deep NGS HLA typing and TCR repertoires were generated after DNA extraction and genomic library preparation.

An healthy control cohort was assembled from three different populations: I) 20 healthy subjects received an NGS HLA typing at Cleveland Clinic and gave their consent to participate to this study; 310 subjects with a known 6 loci HLA genotype were selected from the Allele frequency net database (AFND, USA, San Diego population, gold-standard data classification);^63^ 630 HLA genotypes derived from Emerson and De Witt study.^64^ This last source provided also the control TCR repertoire dataset. The ethnic composition of the control cohort as well as relative HED distributions are reported in supplementary appendix (**Figure S10**).

HLA genotypes of patients with myeloid neoplasia were obtained from the Cleveland Clinic bone marrow transplant registry. T1D dataset was built after HLA allele imputation of data generated from a single nucleotide polymorphism (SNP) array platform and was provided by the Type 1 diabetes genomic consortium (T1DGC, National Institute of Diabetes and Digestive and Kidney Diseases -NIDDK).65,66,67

HLA dataset for the Finnish cohort was provided by the Hematology Research Unit Helsinki in collaboration with the Histocompability Testing Laboratory, Finnish Red Cross Blood Service (N=37 IAA patients and N=42 healthy controls). For this cohort, HLA typing was performed with Luminex Bead Array technology complemented with sequence-specific oligonucleotide primed PCR (PCR-SSO). The bead array data were interpreted according to the manufacturer’s recommendations using the HLA Fusion software 3.2 (One Lambda). The rest of the control cohort was built with genotypes of Finnish individuals extracted from the 1000 Genome project (N=87).^68^

### DNA isolation

Genomic DNA was isolated directly from cryopreserved unfractionated peripheral blood mononuclear cells with the Nuclei Lysis Solution (Promega) according to manufacturer’s instructions.

### HLA sequencing, typing and SNP imputation

A deep NGS panel (TruSight HLA v2 Illumina/Gendex) was used to provide an unambiguous phase-resolved HLA typing for IAA/PNH cohort and 20 healthy subjects from the institutional enrollment. In brief 11 HLA loci (Class I HLA-A, B, and C; Class II HLA-DRB1/3/4/5, HLA-DQA1, HLA-DQB1, HLA-DPA1, and HLA-DPB1) were amplified with a long-range polymerase chain reaction (PCR). Briefly, after amplification a transposone-based DNA tagmentation was applied to generate DNA amplicons, via DNA fragmentation and addition of adapter sequences. Additional PCR steps provided sequence adapters and indexing primers to generate sequencing-ready DNA libraries. Prepared libraries were then loaded directly onto a MiSeq System for sequencing.

High resolution six-digit HLA typing was then inferred with HLA-HD v 1.3,^69,70^ based on the nomenclature of the IPD-IMGT-HLA database v. 3.40.^71^ This tool previously showed its ability to provide an highly accurate HLA inference (miscall rate <4%) in independent studies^72,73^ and was cross-validated in our cohort, for patients with available SSO-PCR HLA typing (**Table S14**).

SNP2HLA v.1.0 was used to impute HLA alleles and SNPs in HLA region from the genotype data provided by the T1DGC as previously described.^74^ For this cohort genotyping was performed using the custom high-density genotyping array Immunochip (Illumina), designed to densely genotype immune-mediated disease loci identified by common variant genome wide association studies (GWAS). With this recommended method, HLA imputation from the Immunochip platform, has been shown to have an accuracy at four-digit resolution between 0.95 and 0.99 across loci.^34^SSO-PCR methods were used to generate HLA genotypes from the myeloid neoplasia cohort in our Institution and from the Finnish cohort in the Helsinki University Hospital.

### Association study and allele risk analysis

High quality 4-digit HLA data, in patient and control cohorts were used for the phenotypic association study and the risk allele analysis. First, frequency of each allele present in IAA/PNH and HC populations was computed by direct counting. The significance of associations with class assignment (phenotype status) was assessed by performing two-tailed Fisher’s exact test applying hypergeometric distribution in a 2 x 2 contingency table. Estimated effect size (odd ratio) was calculated for each allele taking into account either allelic frequencies (additive model) or genotypic frequencies (dominant model), **Tables S1 and S2**.

Given a p-value<0.05, the false discovery rate (FDR) was determined based on the correction for multiple testing (q-value), according to Benjamini and Hochberg procedure.^45^ Alleles identified as differentially distributed between IAA/PNH and HC groups (q-value<0.05) with this procedure were considered for logistic regression analysis to confirm the phenotypic associations. A multivariable logistic regression analysis was applied to test the independent contribution of HED and risk allele profiles on BMF phenotype.

### HED computation

HED scores were computed for all the subjects and all the genotypes in study, using the algorithm published by Pierini and Lenz, applying a customized perl script (https://sourceforge.net/projects/granthamdist/) for the calculation of the amino acid sequence divergence.^41,7^ Briefly, starting from a dictionary including all the protein sequences of exons 2 and 3 for class I alleles and exon 2 for class II alleles, assembled from the IPD-IMGT/HLA database v.3.40^71^,we calculated HED for 6 class I (A, B, C) and II HLA loci (DRB1, DQB1, DPB1).

Means of class I and II were used as quantitative parameters for the analysis of the impact of HED on phenotypic characteristics and clinical outcomes. When appropriate, categorization of high and low intra-locus or intra-class HED was defined according to the 50th percentile of the respective metrics in healthy controls.

### Immunopeptidomic analysis

We used for this analysis a HSC specific proteomic reference as published by Henrich et al.^51^ Amino acids sequences of all the possible alternative transcripts identified based on this protein ID list were selected from the human peptidome reference, downloaded from Ensembl ^75^ (ftp://ftp.ensembl.org/pub/grch37/update/fasta/homo_sapiens/pep//Homo_sapiens.GRCh37.pep.all.fa).

The resulting FASTA file was submitted to NetMHCIIpan 4.0^76^ within a high performant computational environment, in order to determine the binding predictions for selected class I and II HLA molecules for all the peptides with a percentile rank of eluted ligand prediction score <2% for strong binders and <10% for weak binders. This analysis was performed for all the alleles in DRB1 locus and for the most frequent DQB1/DQA1 haplotypes present in our patient and healthy control population.

### Crystallographic structures and molecular dynamics

Crystallographic structures of DR molecules were prepared with the PyMOL (www.pymol.org) program using the known structure of the complex DRA*01:01-DRB1*15:01-MBP (PDB: 1BX2,^77^ resolution: 2.6 Å), DRB1*14:02-DRA*01:01/ vimentin (amino acid positions: 59-71, PDB:1H15^78^, resolution: 3.1 Å) and DRB5*01:01-DRA*01:01/EBV DNA polymerase (628-641) (PDB:6ATF^79^, resolution: 1.9 Å). Homology models of DRB1*16:01 and DRB1*12:01 were constructed using the software I-TASSER,^80,81^ and the protein sequences of DRB1*16:01:01 and DRB1*12:01:01:01 alleles were extracted from the IPD-IMGT/HLA database. All model structures were superimposed to the known structure of DRB1*15:01. DRA*01:01:01:01 was used as reference for the α chain in the three structures. Model structures of HLA with MBP were subject to molecular dynamic (MD) simulations and analyses using the Amber18 program.^82^ The protocol of MDsimulations was conducted according to a previous study.^83^ Thirty-six ns production-run MD simulations were performed to calculate the fluctuations of the antigen MBP within the DR binding groove (as in **Fig S11**). Twenty conformations in the 16-36 ns MD simulations were used to calculate the total binding free energy between HLA and MBP using the MM-GBSA method.^84^ The TCR α and β chains, DRA*01:01-DRB1*15:01 and CD4 model structure was constructed by aligning HLA/peptide with the peptide-MHC in the crystal structure of TCR, peptide-MHC, CD4 (PDB: 3T0E).

### TCRβ chain sequencing and analysis

Immunosequencing of the CDR3 regions of human TCRβ chains was performed using the ImmunoSEQ Assay (Adaptive Biotechnologies, Seattle, WA), as previously described. ^85,86,87^ In brief, extracted genomic DNA was amplified in a bias-controlled multiplex PCR, with i) a first PCR step consisting in forward and reverse amplification primers specific for every V and J gene segment, to allow the amplification of the hypervariable CDR3 region, and ii) a second PCR adding a proprietary barcode sequence and Illumina adapter sequences. CDR3 libraries were sequenced on an Illumina MiSeq system according to the manufacturer’s instructions. Rearrangement details from healthy controls (HC, age-matched with donors recruited in our cohort) were derived from the Emerson and DeWitt study (data provided by the original publication and the ImmuneACCESS platform).^64,88^ ImmunoSeq Analizer 3.0 suite was used for sample export and preliminary statistics and quality control steps while R bioconductor^89^ environment and Immunarch R^90^ suite were used for all the downstream analyses (see supplementary appendix).

All metrics were calculated based only on the “productive” rearrangements (translating a functional amino acids sequence, intended as templates that were in-frame and did not contain a stop codon in their sequence) within the normalized down-sampled dataset. Details concerning the bioanalytic workflow are reported in the supplementary appendix. Sequencing results are accessible through the publicly available repository of ImmuneAccess platform.

### Statistical analysis

All metrics in study were generally treated as continuous variables and categorized when needed. Median, interquartile ranges (IQR), mean and 95%CI intervals were used where appropriate. Frequency and distribution of categorical variables were expressed as percentage. For all relevant comparisons, after testing for normal distribution, comparative analyses between two groups were performed, by two-sided paired or unpaired Student’s t-tests at 95% CI. In the case of not normally distributed data, the wilcoxon matched-pairs signed rank test at 95% CI was used. Fisher’s exact test or Chi-square were applied for independent group comparisons, in case of testing more than two groups a one-way ANOVA test was used. Correction for multiple testing was made through Benjamini and Hochberg correction.^45^ Cox regression and proportional hazard models for the competing risk sub distributions were used to assess the impact of HED metrics on clinical outcomes (OS, cumulative incidence of progression) in univariable setting.^91^ All statistical tests were two-sided, and a P-value <0.05 was considered statistically significant. OS was defined as the time from diagnosis to the last follow-up or death for any cause. Death at any time was used as competitive events for the cumulative incidence of progression to MDS. All of the analyses and data visualization were performed using the statistical computing environment R (4.0.0 R Core Team, R Foundation for Statistical Computing, Vienna, Austria) and excel Microsoft 365.

## Data Availability

All the data that support the findings of this study are available within the Article and Supplementary Files. Genotypic and phenotypic raw data, including HLA genotypes of all the subject in study will be provided upon request to the corresponding authors. TCR sequencing data will be provided through the ImmuneACCESS platform (Adaptive Biotechnology).

## AUTHORSHIP AND DISCLOSURES

### Authorship contributions

SP designed the study, collected, analyzed and interpreted the data, performed the bioinformatic and statistical analyses and wrote the manuscript. CG performed NGS experiments, clinical data collection and participated in the analysis interpretation. SK, LT, MZ, AK, HA, YG, BJK helped in sample and data collection. TLa and VV edited the manuscript, helped in data interpretation and gave helpful intellectual insights during the study. YS, BJP, MN, BH actively participated in patient recruitment, management, and follow-up. SL and SM provided genotypic data for the Finnish cohort and critically revised the manuscript. CY performed the molecular modeling, analyzed the crystallographic structures helped in drafting the methods inherent to this part. TC and TLe helped in data interpretation and analytical method development. JM designed and conceptualized the study, interpreted the data analysis and edited the manuscript. First and last authors took responsibility for the integrity and the accuracy of the data presented. All authors reviewed and approved the final version of this manuscript.

## Acknowledgements

This work was supported by US National Institute of Health (NIH) grants R35 HL135795, R01HL123904, R01 380HL118281, R01 HL128425, R01 HL132071, Edward P.Evans Foundation (to J.M), Italian Society of Hematology, Fondation ARC pour la Recherche sur le Cancer, Philippe Foundation, Association HPN France / Fondation maladies rares (to S.P.), The American-Italian Cancer Foundation (to C.G.); VeloSano Pilot Award, and Vera and Joseph Dresner Foundation– MDS (to V.V.). European Research Council (M-IMM and STRATIFY projects), Academy of Finland, Sigrid Juselius Foundation, and Cancer Foundation Finland (to S.L. and S.M).We thank Diego Chowell for his helpful insights and critical revision of the manuscript.

We thank the National Institute of Diabetes and Digestive and Kidney diseases and the Type 1 Diabetes Genetic Consortium that provided the T1DGC Immunochip/HLA Reference Panel used in this study and Dr. John Sidney and Prof. Alessandro Sette who kindly provided the ethnicity/race information for the subjects from San Diego population.

