## Supplementary Appendix for "The Immunogenetic Basis of Idiopathic Bone Marrow Failure Syndromes: A Paradox of Similarity and Self-Presentation"

#### Description of supplemental material:

All the supplementary tables are provided as separate Excel files.

- Supplemental table 1. Frequency and association analysis of HLA alleles (Genotypic frequencies; Dominant effect)
- Supplemental table 2. Frequency and association analysis of HLA alleles (Allelic frequencies; Additive effect)
- Supplemental table 3: Genotypic frequency and distribution of risk alleles in myeloid neoplasia and control group (Dominant effect)
- Supplemental Table 4: Univariable logistic regression analysis of risk allele associations with "enriched" immune-mediated phenotypes
- Supplemental Table 5: Univariable logistic regression analysis of class II HED associations with "enriched" immune-mediated phenotypes
- Supplemental Table 6: HED scores across all the cohorts in study
- Supplemental Table 7**:** Simulated divergences between DRB1*15:01 and other DRB1 alleles
- Supplemental Table 8: Simulated divergences between DQB1*06:02 and other DQB1 alleles
- Supplemental Table 9: Recursive analysis of the variable aminoacidic the region of antigen binding site of DRB1 locus
- Supplemental Table 10: Recursive analysis of the variable amino acidic region of antigen binding site of DQB1 locus
- Supplemental Table 11: Number of total binders per protein coding gene for each DRB1 molecule
- Supplemental Table 12: Number of total binders per protein coding gene for each DQA1-DQB1 molecule
- Supplemental Table 13: Race/Ethnicity distribution of the North American healthy control and patient populations
- Supplemental Table 14: Race stratified distribution or risk alleles in the populations in study
- Supplemental Table 15: Internal validation of NGS based typing with HLA-HD program

##### Supplemental Figure Legend

**Figure S1:** Flow chart describing the design and the principal steps of the study.
**Figure S2:** Risk allele association analysis with disease subcategories and binary outcomes. A) Frequency of class I and II risk alleles in idiopathic aplastic anemia (IAA) and primary and secondary paroxysmal nocturnal hemoglobinuria (PNH).
Binomial logistic regression analysis for association between risk alleles and response to immunosuppression (B), progression to myelodysplastic syndrome (MDS)/acute myeloid leukemia (AML), (C); evolution to secondary PNH (D). Each outcome is considered as binomial variable.

**Figure S3:** Distribution of class and locus specific HLA evolutionary divergence (HED) scores in healthy controls (HC) and IAA/PNH according to the presence/absence of class II risk alleles.

Numbers of non-carriers, homozygous and heterozygous subjects are reported for each group.

A) Mean class II HED scores in HC carriers and non-carriers of DRB1*15:01.
B) DRB1 HED scores in HC carriers and non-carriers of DRB1*15:01.
C) Mean class II HED scores in patients carriers and non-carriers of DRB1*15:01.
D) DRB1 HED scores in patients carriers and non-carriers of DRB1*15:01.
E) Mean class II HED scores in HC carriers and non-carriers of DQB1*06:02.
F) DQB1 HED scores in HC carriers and non-carriers of DQB1*06:02.
G) Mean class II HED scores in patients carriers and non-carriers of DQB1*06:02.
H) DQB1 HED scores in patients carriers and non-carriers of DQB1*06:02.

**Figure S4:** Distribution of mean class II and DRB1 HED scores for each cohort in study.
HC: healthy controls; IAA/PNH: Idiopathic aplastic anemia/paroxysmal nocturnal hemoglobinuria; MN: myeloid neoplasia; T1D: Type 1 diabetes.

A) Violin plots showing the distribution of mean class II HED scores in all the cohorts (Wilcoxon signed rank test).
B) Violin plots showing the distribution of DRB1 HED scores in all the cohorts (Wilcoxon signed rank test).
C) Proportion of homozygosis and heterozygosis of DRB1 locus in all the cohorts in study.
D) Univariable logistic regression analysis predicting the association between DRB1-related predictors and disease phenotype (comparisons with HC group) .

**Figure S5**: Analysis of associations between class-related HED scores and characteristics and outcomes of IAA/PNH patients

A) Linear regression analysis between class I HED and granulocytic PNH clonal size at diagnosis.
B) Linear regression analysis between class II HED and granulocytic PNH clonal size at diagnosis.
C) Linear regression analysis between class I HED and age at diagnosis.
D) Linear regression analysis between class II HED and age at diagnosis.
E) Mean class I HED scores in HC (dotted red line indicates the $50^{th}$ percentile used as cutoff for categorization in high and low HED in disease population).
F) Mean class II HED scores in HC (dotted red line indicates the $50^{th}$ percentile used as cutoff for categorization in high and low HED in disease population).
G) Univariable analysis of impact of mean class I HED on probability of survival. Hazard ratio calculated by applying cox regression proportional models. P-value resulting from the log-rank test between the two time dependent probability distributions. Primary PNH patients were excluded from this analysis.
H) Univariable analysis of impact of mean class II HED on probability of survival. Hazard ratio calculated by applying cox regression proportional models. P-value resulting from the log-rank test between the two time dependent probability distributions. Primary PNH patients were excluded from this analysis.
I) Univariable analysis of impact of mean class I HED on cumulative incidence of malignant progression. Hazard ratio calculated by applying proportional subdistribution of hazards, accommodating competing risk of death.
J) Univariable analysis of impact of mean class II HED on cumulative incidence of malignant progression. Hazard ratio calculated by applying proportional subdistribution of hazards, accommodating competing risk of death.

K) Univariable analysis of impact of mean class I HED on non-malignant progression towards secondary PNH. Hazard ratio calculated by applying proportional subdistribution of hazards, accommodating competing risk of death. Primary PNH patients were excluded from this analysis.
L) Univariable analysis of impact of mean class II HED on non-malignant progression towards secondary PNH. Hazard ratio calculated by applying proportional subdistribution of hazards, accommodating competing risk of death. Primary PNH patients were excluded from this analysis.

Hazard ratio in panels H,J,L denotes the risk that low mean class II HED confers to the outcomes in analysis.

**Figure S6: Recursive analysis of the amino acid sequence within the peptide binding site of DQB1 locus**

A) Lower panel: WebLogo visualization representing the contribution of single amino acids within the variable portion of the peptide binding site of DRB1 locus (https://weblogo.berkeley.edu/). The x-axis indicates each variable position (as per IPD-IMGT-HLA reference). Letters represent each possible amino acid at each given position; Letters' height illustrates the frequency of each amino acid in healthy control population. Colors indicate the chemico-physical properties as per legend in B.
B) Upper panel: stylized visualization of amino acids enriched in IAA/PNH cohort. Letters' height illustrates the frequency of each amino acid in disease population. Markers indicate the odd ratios resulting from the logistic regression analysis studying the amino acids contribution in determining disease phenotype (see methods and Table S8).
C) Scatter plot showing the significance level of each variable amino acid in the peptide binding site of DRB1 locus found enriched in IAA/PNH population compared to HC. Each dot represents the negative logarithm base 10 for the adjusted p-value (q-value/FDR) referring to each amino acid. The position on x-axis indicates the position within the peptide binding site according to IPD-IMGT-HLA reference. Only the alleles including the amino acids enriched in IAA/PNH group are indicated in the boxes. Colors represent the chemico-physical properties as per legend.

**Figure S7: Hematopoietic stem cell specific immune peptidomic binding capacities of the DQ molecules.**

A) Filled 2D area plot representing the binding capacities of each DQA1-DQB1 molecule in study for the self-peptidomic HSC reference. Y-axis indicates the percentage (%) of self-peptides predicted to bind DRB1 molecules. Red portion indicates strong binders, pink portion of the graph depicts instead weak binders (see methods). The pie chart indicates the proportion of binders and non-binders for the whole reference.
B) Number of strong (red) and weak binders (pink) bound by different DQB1*06:02 molecules.
C) Heatmap showing the top-100 coding genes associated with the self-peptidome formation. Each cell represents the Z-score of the number of binders predicted of each transcript for a given DQ molecule.
D) Distribution of the number of binders predicted for DQ molecules in HC and IAA/PNH groups. Wilcoxon signed rank test.

**Figure S8: Validation of DRB1-related immunogenetic determinants in a uniquely HLA assorted population**

A) Differences in distribution of DRB1*15:01 between Finnish patients and ethnicity-matched HCs. Chi square.
B) Differences in distribution of homozygous alleles in DRB1 locus between Finnish patients and ethnicity-matched HCs. Chi square.
C) Distribution of DRB1 HED scores in Finnish IAA and ethnicity-matched HC subjects (Wilcoxon signed rank test).
D) Violin plot showing the distribution of DRB1 HED scores in Finnish IAA patients (N=30) and ethnicity-matched HCs (N=128) (Wilcoxon signed rank test).
E) Violin plot showing the distribution of DRB1 HED scores in Finnish non DRB1*15:01 carriers [ IAA patients (N=11) and HCs (N=96), Wilcoxon signed rank test].
F) Violin plot showing the binding capacities of DRB1 alleles in Finnish IAA patients (N=30) and ethnicity-matched HCs (N=128).

**Figure S9: T cell receptor analysis in IAA patients**

A) Inverse simpson index (ISI) distribution in HC and IAA patients (down-sampled dataset). Violin plots showing median and interquartile ranges. Wilcoxon signed rank test.
B) Number of unique clonotypes in HC and IAA patients (down-sampled dataset). Violin plots showing median and interquartile ranges. Wilcoxon signed rank test.
C) Mean size of expansion of each clonotype of size $\geq$2 templates in HC and IAA patients (down-sampled dataset). Violin plots showing median and interquartile ranges. Wilcoxon signed rank test.
D) ISI distribution in HC and IAA patients in DRB1*15:01 carriers (down-sampled dataset). Violin plots showing median and interquartile ranges. Wilcoxon signed rank test.
E) ISI distribution in HC and IAA patients in non-DRB1*15:01 carriers (down-sampled dataset). Violin plots showing median and interquartile ranges. Wilcoxon signed rank test.
F) Linear regression analysis between ISI and mean class I HED.
G) Linear regression analysis between number of unique clonotypes and mean class I HED
H) Linear regression analysis between mean size of clonotype expansion and mean class I HED.
I) Linear regression analysis between ISI and mean class II HED.
J) Linear regression analysis between number of unique clonotypes and mean class II HED.
K) Linear regression analysis between mean size of clonotype expansion and mean class II HED.
R-squared goodness-of-fit are reported along with the p-value in each box (from F to K).
L) Proportion of known complementary determining region 3 (CDR3) specificities in IAA and HC groups (this distribution has been computed in the downsampled dataset).
M) Negative logarithm of mean frequency of autoreactive clonotypes present in HC and IAA patients. Violin plots showing median and interquartile ranges. Each dot represent the mean frequency/per subject (All the values refer to the non-downsampled dataset in order to capture all the possible recognizable CDR3 sequences). Wilcoxon signed rank test
N) Linear regression analysis between frequency of known autoreactive clonotypes in IAA patients and mean class II HED. Each dot represents one autoreactive clonotype. Clonotypes with overlapping frequencies are represented by darkest dots. (All the values refer to the non-downsampled dataset). Wilcoxon signed rank test. R-squared goodness-of-fit are reported along the p-value.

O) Bubble matrix showing the mean frequency of each autoimmune-disease associated clonotype present in the non-downsampled repertoires of IAA and HCs. Each bubble represents the number of clonotypes with known autoimmune specificity (X-axis). The size of each bubble indicates the mean frequency. Wilcoxon signed rank test is used to compare the mean frequencies of each specificity between IAA and HC groups.

**Figure S10:** Race distribution in healthy controls and aplastic anemia patients

(**Hispanic or Latino* category has been integrated in this classification although properly referring to an ethnicity class)

A) Race distribution in HC.
B) Race distribution in IAA/PNH patients.
C) Allelic frequencies in HC and IAA/PNH patients in Caucasian subgroup. P-values calculated with Fisher Exact test on 2x2 contingency tables. Only significant values are shown. * 0.05-0.01; ** <0.01; *** <0.001; ****<0.0001; ***** <0.00001. Allelic frequency computation. Allelic frequencies are calculated as following: # of time each allele is present in the population/ number of haplotypes profiled for the given locus.
D) HED distributions in North American HC population used in our study.
E) Mean class I and class II HED comparisons between HC and IAA/PNH within the Caucasian subset.

**Figure S11:** Molecular dynamic simulations of DR-antigen complexes

**A)** Six representative conformations of DRB1*15:01-DRA*01:01 complex bound to Myelin basic protein (MBP) from 36 ns molecular dynamic (MD) simulations based on the crystal structure (PDB: 1BX2).
**B)** Six representative conformations of DRB1*16:01-DRA*01:01 complex bound to MBP from 36 ns MD simulations. The structure of DRB1*16:01 was generated from the homology modeling using the I-TASSER program.
**C)** Six representative conformations of DRB1*12:01-DRA*01:01 complex bound to MBP from 36 ns MD simulations. The structure of DRB1*12:01 was generated from the homology modeling using the I-TASSER program.
Numbers within the blue line indicate the total binding free energy calculated using the MM-GBSA method using 20 conformations of the 36 ns MD simulations. The lower value indicates a higher affinity between the HLA and the MBP peptide. MBP binds to DRB1*1501 more potently, followed by DRB1*1601 and DRB1*1201.

### Supplemental methods

#### IBMF study cohort

Two-hundred-sixty-three patients diagnosed with idiopathic aplastic anemia (IAA) with or without paroxysmal nocturnal hemoglobinuria (PNH) clone or with primary hemolytic PNH and were included in this study. Diagnosis of IAA was conducted according to established criteria[1],[2] including hypocellular bone marrow (<30% cellularity), without evidences of fibrosis or malignant cells, and at least two of the following cytopenias: i) absolute neutrophil counts (ANC) <1.5 × $10^9$/L (moderate IAA), <0.5 x $10^9$/L (severe) or <0.2 x $10^9$/L (very severe IAA); ii) platelet counts <150 × $10^9$/L (moderate IAA) or <20 x $10^9$/L (severe or very severe IAA); iii) absolute reticulocyte counts (ARC) <60 x $10^9$/L. Diepoxybutane testing was performed for all young adult patients suspected for Fanconi anaemia, and other forms of congenital bone marrow failure were excluded by appropriate testing depending on the clinical scenario. All patients were periodically evaluated for clonal evolution to PNH or myeloid malignancies, defined as any of the following according to WHO 2016 criteria: diagnosis of AML; diagnosis of MDS.[3]

PNH diagnosis was based on the deficiency of GPI-anchored protein on multiple cell lines by flow cytometry and PIGA gene sequencing, as previously shown.[4] Criteria of classical hemolytic PNH were PNH granulocytic clone size >20% and lactate dehydrogenase >x2.5 upper limit of normality.

#### PNH flow cytometry

Cells were acquired on a FC500 or XL-MCL (Beckman Coulter). A 5-color cocktail (CD15-V450, CD45-PC7, CD64-APC, CD157-PE, FLAER-Alexa 488) to identify granulocytes with GPI-linked protein deficiency was deployed to determine PNH granulocytes clone size while CD59-PE (Invitrogen, MHCD5904) and CD235a-FITC (Beckman Coulter (BC), IM2212U) staining was used for determination of PNH red blood cells.[5,6,7]

#### Bioanalytic workflow for TCR analysis

Raw sequences were demultiplexed according to Adaptive's proprietary barcode sequences. Demultiplexed reads were then further processed to: remove adapter and primer sequences; identify and correct for technical errors introduced through PCR and sequencing. The data were filtered and clustered using a modified nearest-neighbor algorithm, to merge closely related sequences.

The resulting sequences were analyzed trough the ImmunoSeq Analizer 3.0 suite which allowed the annotation of the VDJ genes constituting each unique CDR3 and the translation of the encoded CDR3 amino acid sequence. V, D and J gene definitions were based on annotation in accordance with the IMGT database (www.imgt.org).

After sample export (sample overview and rearrangement details), downstream analyses were conducted with the Immunomind/immunarch v.0.6.6 R suite,≈ and the R Bioconductor environment.[8] To overcome the issue related to differences in inter-sample depth, we performed a normalization procedure, resampling the immune repertoire for all the specimens, to a depth of 5240 templates (smallest sample depth in study). The aforementioned tool was used for this procedure setting the following parameters: [*Immunarch function: repSample, parameters: .method = "downsample", .prob = TRUE*].

Diversity was characterized computing:

- The number of all unique clonotypes (richness)
- The size of each clonotypes (evenness or relative abundance).
- The Inverse Simpson`s index (a metrics derived from ecology and used to characterize the alpha diversity),[9,10] calculated according to the following formula:

$$\frac{1}{\lambda} = \frac{1}{\sum_{i=1}^{R} p_i^2}$$

  where $p_i$ is the proportional abundance for each clonotype and *R* is the total number of clonotypes in the sample.

Amino acidic CDR3 sequences present in samples in study were annotated through the integration of a dataset aggregating all public databases supported by the Adaptive Immune Receptor Repertoire Community project. Briefly, after filtering for human TRB data, we downloaded the following public databasets: VDJDB (https://vdjdb.cdr3.net/search), McPAS-TCR (http://friedmanlab. weizmann.ac.il/McPAS-TCR/) and PIRD TBAdb (https://db.cngb.org/pird/tbadb/). Immunarch *dbAnnotate* and *trackClonotypes* functions were used respectively to annotate and track TCR immune specificities.

#### Supplementary considerations

This study aims at the identification of the immunogenetic patterns associated with idiopathic aplastic anemia susceptibility. Here we comprehensively assess the structures involved in antigen presentation in a cohort of 263 IAA North American subjects, encompassing risk allele analysis, functional and structural HLA evolutionary divergence, binding prediction and molecular modelling. We also analysed T cell receptor (TCR) clonotypic interactions and related specificities.

The choice to use a mixed population both in control and in disease groups arises from the need to identify superstructures embracing race and ethnic background, with at the same time, a statistical power adequate to the sample size of a rare disease. With that in mind, although our main analyses were addressed to the entire population in study, different subanalyses were performed on race/ethnicity stratified subgroups as well as on a Caucasian-only population (**Supp. Table 13 and Fig. S10**). Specifically, since with the two genetic models applied on the entire cohort, we were not able to identify any of the recently described class I risk alleles (B*14:02 and B*40:02)[15,16] . Allelic frequency analysis was also carried out on the Caucasian subset only. Indeed, after confirming the alleles identified through our main analysis, we found a small but significant increase in frequency for B*14:02 (4.7% in IAA vs 1.9% in HC, p=0.002) but not for B*40:02 (0.74% vs 2.05% respectively, p=0.111). Other alleles unveiled in this subanalysis were: C*08:02 (5.3% vs 2.4%, p= 0.0080) and DRB1*01:02 (3.3% vs 1.2%, p= 0.00002). Nevertheless, the fraction of IAA cases attributable to these alleles was very low in our study with only a few carriers accounting for the nominal difference in the frequency between patients and controls, restricting the potential etiologic fraction associated with disease phenotype.

More generally speaking, the relative risk conveyed by the presence of susceptibility HLA alleles, given the rarity of this condition, is relatively low, and "contamination" of experimental cohorts with non-immune mediated cases remain possible despite application of stringent diagnostic criteria. Therefore, we demonstrated the enrichment in susceptibility profiles also when analyzing subsets of patients responding to immunosuppression and harboring a PNH clone as surrogate markers of immune-mediated IAA cases, with stronger associations demonstrated for DRB1 and DQB1 risk alleles (**Table S4**).

The structural differences between patients and controls in terms of functional divergence of class II loci, with reproducibility in ethnicity/population restricted subcohorts and in different autoimmune disorders (**Fig. S4, S8, S10**), emphasize the role of these antigen-presenting structures in generating the autoimmune proclivity, eventually hampering the homeostatic control of autoreactive T cell clones.

How much this environment, defining prevalently an aberrant CD4+-restricted immunity, may debilitate also CD8+ responses, representing instead the operative arm of the hematopoietic stem cell destruction in IAA, remains to be elucidated. The use of quantitative metrics such as HLA evolutionary divergence or peptide binding predictions and their impact on adaptive immune perturbations may definitely help shedding light on the pathophysiology involved in organ-specific autoimmunity and in the central tolerance processes of all immune-mediated disorders.

**Figure S1**

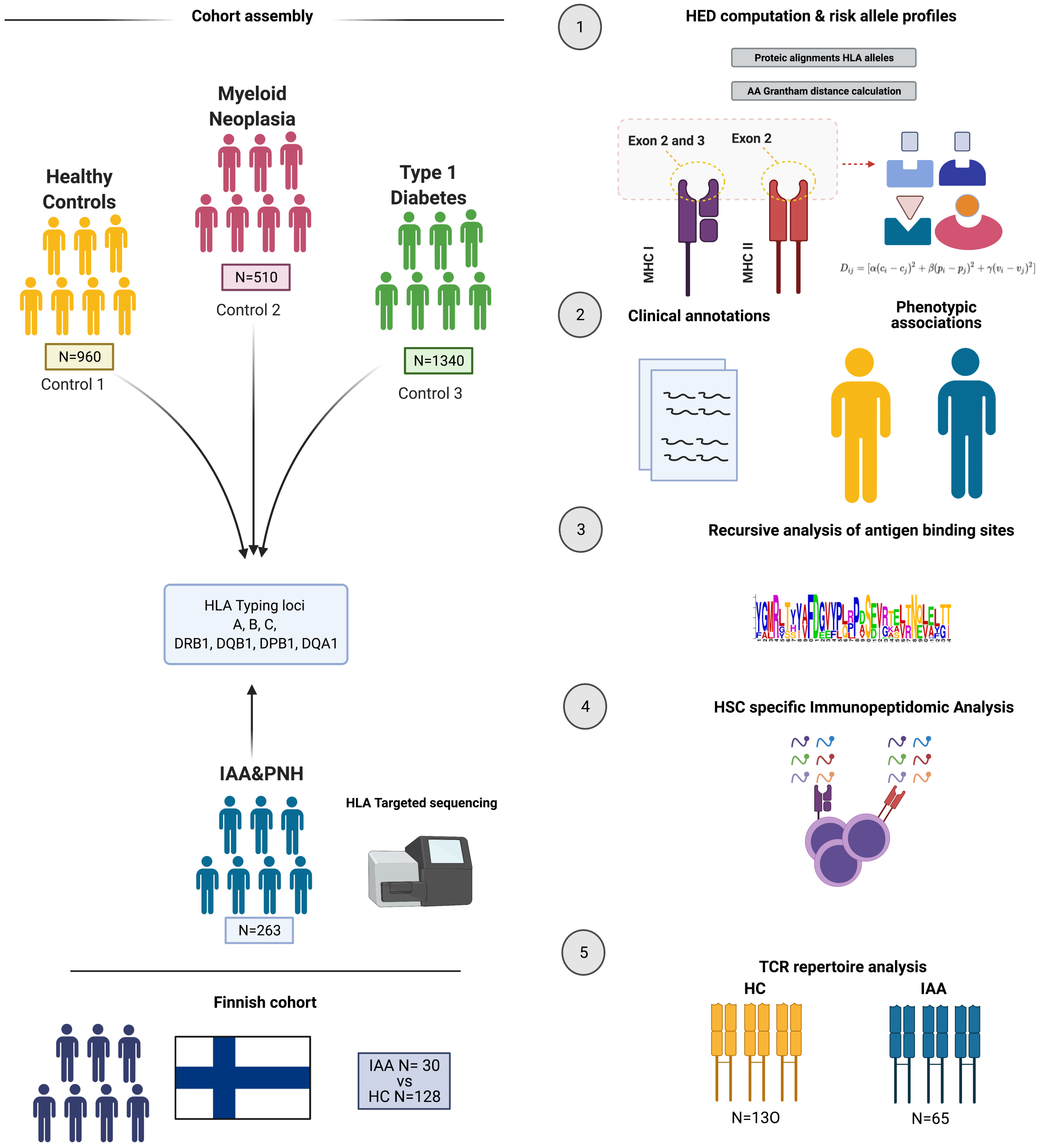
Cohort assembly
Myeloid Neoplasia
N=510
Control 2
Healthy Controls
N=960
Control 1
Type 1 Diabetes
N=1340
Control 3
HLA Typing loci
A, B, C,
DRB1, DQB1, DPB1, DQA1
IAA&PNH
N=263
HLA Targeted sequencing
Finnish cohort
IAA N= 30
vs
HC N=128
1
HED computation & risk allele profiles
Proteic alignments HLA alleles
AA Grantham distance calculation
Exon 2 and 3
Exon 2
MHC I
MHC II
$D_{ij} = [\alpha(c_i - c_j)^2 + \beta(p_i - p_j)^2 + \gamma(v_i - v_j)^2]$
2
Clinical annotations
Phenotypic associations
3
Recursive analysis of antigen binding sites
4
HSC specific Immunopeptidomic Analysis
5
TCR repertoire analysis
HC
IAA
N=130
N=65

**Figure S2**

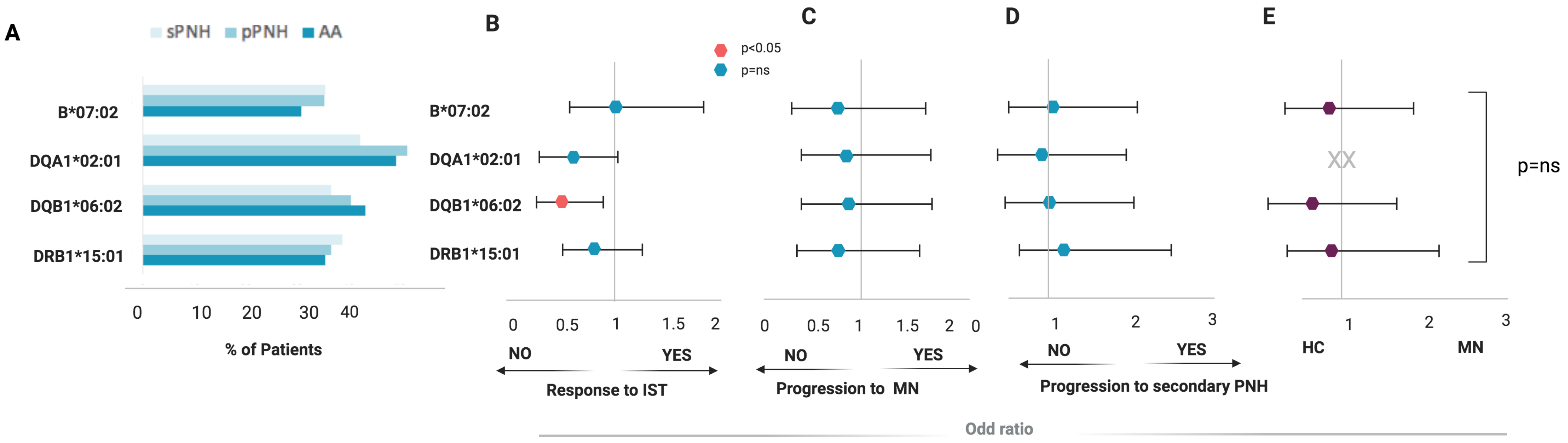
A
sPNH
pPNH
AA
B*07:02
DQA1*02:01
DQB1*06:02
DRB1*15:01
0
10
20
30
40
% of Patients
B
C
D
E
p<0.05
p=ns
B*07:02
DQA1*02:01
DQB1*06:02
DRB1*15:01
0
0.5
1
1.5
2
NO
YES
Response to IST
0
0.5
1
1.5
2
NO
YES
Progression to MN
0
1
2
3
NO
YES
Progression to secondary PNH
XX
1
2
3
HC
MN
p=ns
Odd ratio

**Figure S3**

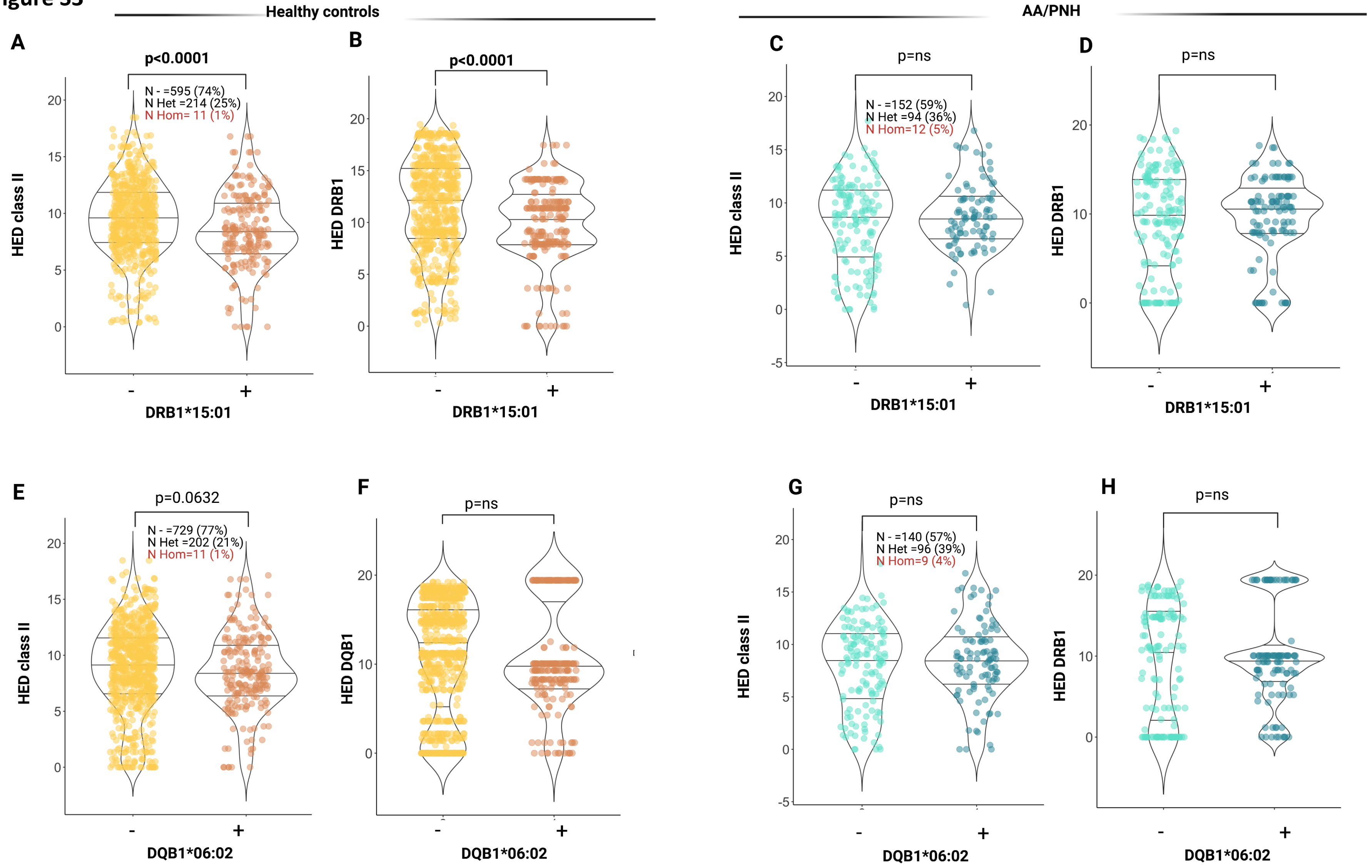
Healthy controls
AA/PNH
A
p<0.0001
N - =595 (74%)
N Het =214 (25%)
N Hom= 11 (1%)
HED class II
DRB1*15:01
B
p<0.0001
HED DRB1
DRB1*15:01
C
p=ns
N - =152 (59%)
N Het =94 (36%)
N Hom=12 (5%)
HED class II
DRB1*15:01
D
p=ns
HED DRB1
DRB1*15:01
E
p=0.0632
N - =729 (77%)
N Het =202 (21%)
N Hom=11 (1%)
HED class II
DQB1*06:02
F
p=ns
HED DQB1
DQB1*06:02
G
p=ns
N - =140 (57%)
N Het =96 (39%)
N Hom=9 (4%)
HED class II
DQB1*06:02
H
p=ns
HED DRB1
DQB1*06:02

#### Figure S4

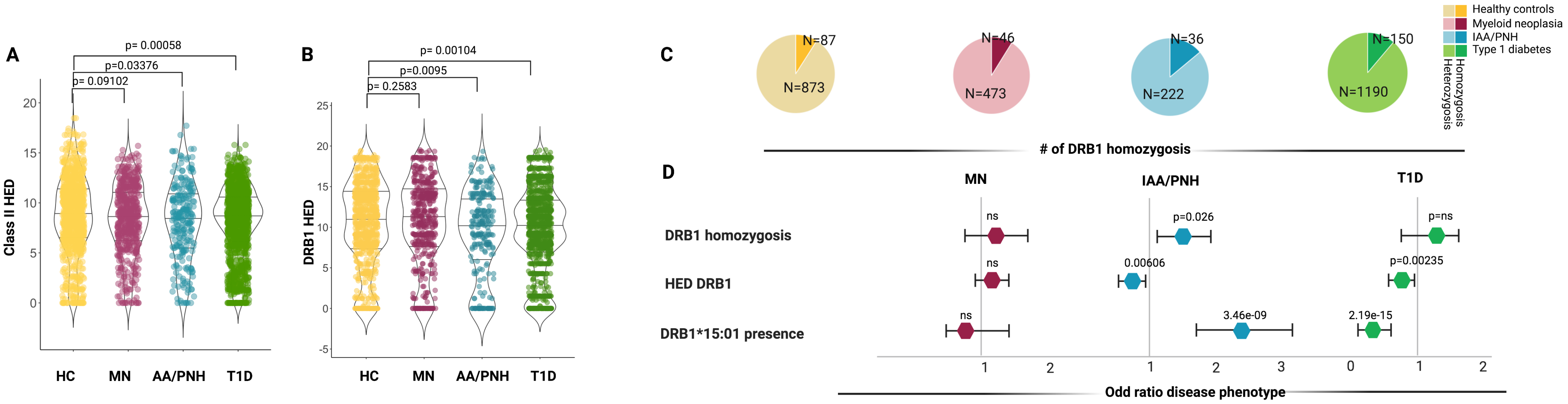
A
p= 0.00058
p=0.03376
p= 0.09102
Class II HED
HC
MN
AA/PNH
T1D
B
p= 0.00104
p=0.0095
p= 0.2583
DRB1 HED
HC
MN
AA/PNH
T1D
C
N=87
N=873
N=46
N=473
N=36
N=222
N=150
N=1190
Healthy controls
Myeloid neoplasia
IAA/PNH
Type 1 diabetes
Homozygosis
Heterozygosis
### of DRB1 homozygosis
D
MN
IAA/PNH
T1D
DRB1 homozygosis
HED DRB1
DRB1*15:01 presence
ns
p=0.026
p=ns
ns
0.00606
p=0.00235
ns
3.46e-09
2.19e-15
Odd ratio disease phenotype

#### Figure S5

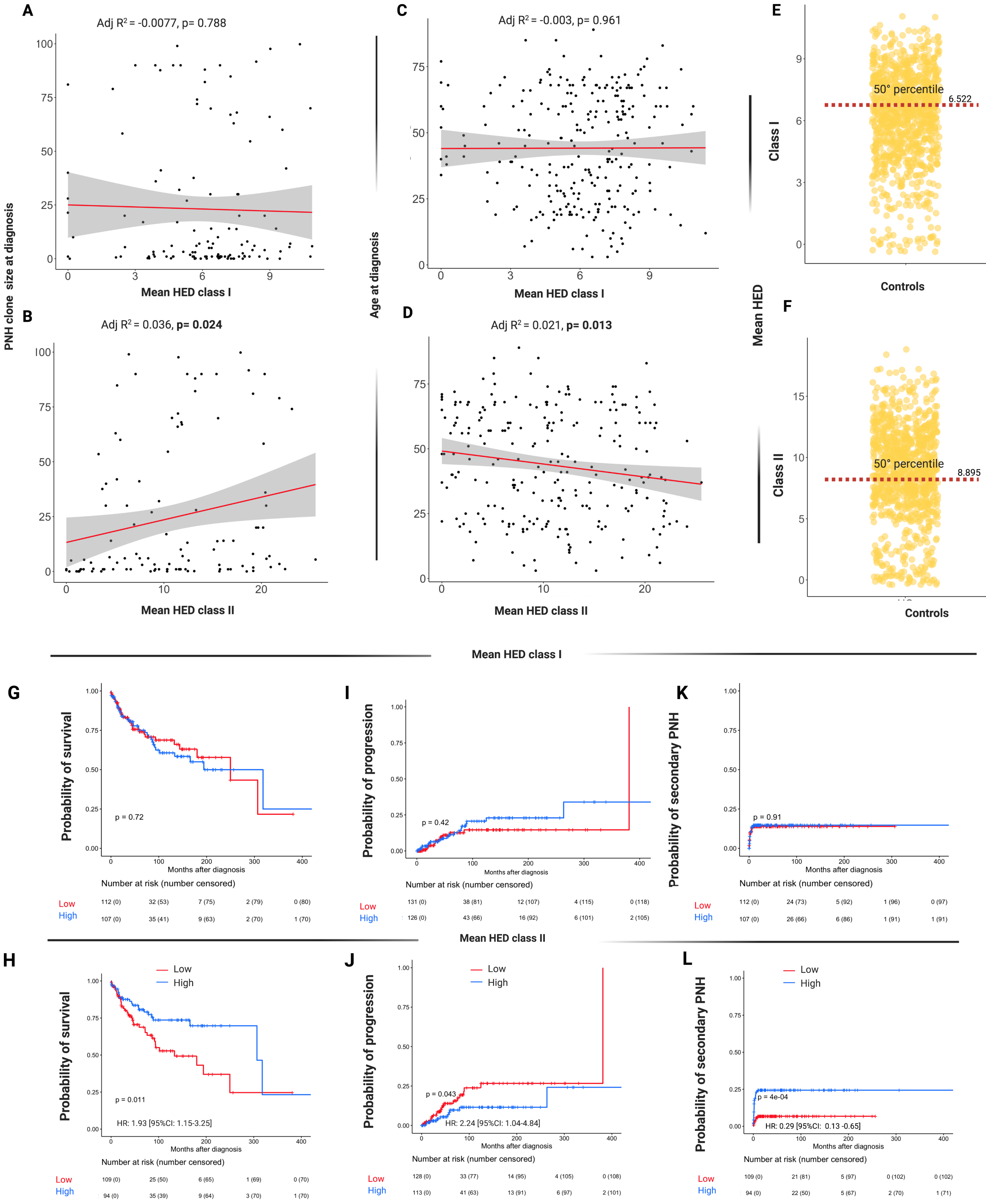
A
Adj R2 = -0.0077, p= 0.788
PNH clone size at diagnosis
Mean HED class I
B
Adj R2 = 0.036, p= 0.024
Mean HED class II
C
Adj R2 = -0.003, p= 0.961
Age at diagnosis
Mean HED class I
D
Adj R2 = 0.021, p= 0.013
Mean HED class II
E
Mean HED
Class I
50° percentile
6.522
Controls
F
Class II
50° percentile
8.895
Controls
Mean HED class I
G
Probability of survival
p = 0.72
Months after diagnosis
Number at risk (number censored)
Low 112 (0) 32 (53) 7 (75) 2 (79) 0 (80)
High 107 (0) 35 (41) 9 (63) 2 (70) 1 (70)
I
Probability of progression
p = 0.42
Months after diagnosis
Number at risk (number censored)
Low 131 (0) 38 (81) 12 (107) 4 (115) 0 (118)
High 126 (0) 43 (66) 16 (92) 6 (101) 2 (105)
K
Probability of secondary PNH
p = 0.91
Months after diagnosis
Number at risk (number censored)
Low 112 (0) 24 (73) 5 (92) 1 (96) 0 (97)
High 107 (0) 26 (66) 6 (86) 1 (91) 1 (91)
Mean HED class II
H
Low
High
Probability of survival
p = 0.011
HR: 1.93 [95%CI: 1.15-3.25]
Months after diagnosis
Number at risk (number censored)
Low 109 (0) 25 (50) 6 (65) 1 (69) 0 (70)
High 94 (0) 35 (39) 9 (64) 3 (70) 1 (70)
J
Low
High
Probability of progression
p = 0.043
HR: 2.24 [95%CI: 1.04-4.84]
Months after diagnosis
Number at risk (number censored)
Low 128 (0) 33 (77) 14 (95) 4 (105) 0 (108)
High 113 (0) 41 (63) 13 (91) 6 (97) 2 (101)
L
Low
High
Probability of secondary PNH
p = 4e-04
HR: 0.29 [95%CI: 0.13 -0.65]
Months after diagnosis
Number at risk (number censored)
Low 109 (0) 21 (81) 5 (97) 0 (102) 0 (102)
High 94 (0) 22 (50) 5 (67) 2 (70) 1 (71)

**Figure S6**

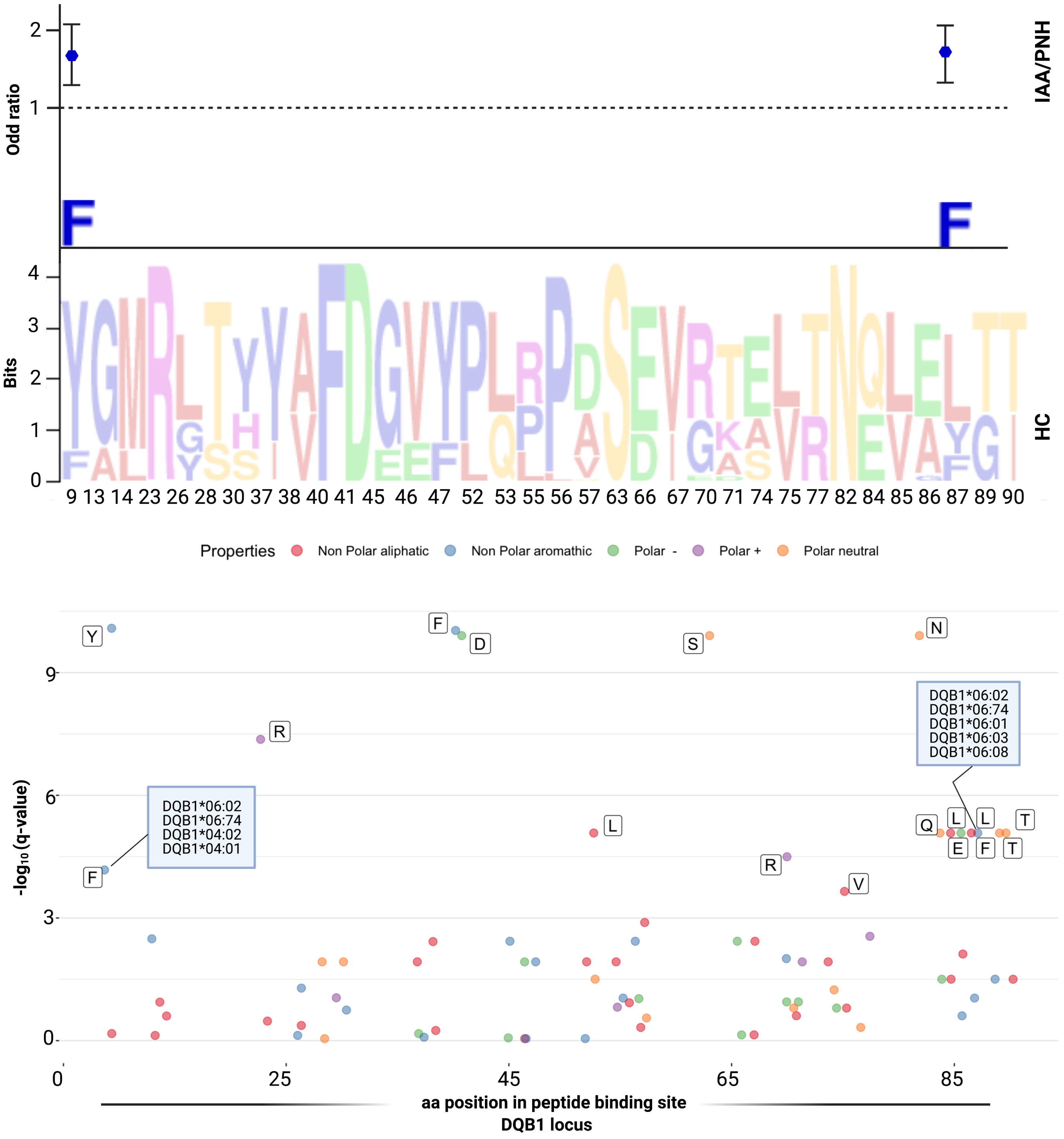
Odd ratio
IAA/PNH
Bits
HC
9 13 14 23 26 28 30 37 38 40 41 45 46 47 52 53 55 56 57 63 66 67 70 71 74 75 77 82 84 85 86 87 89 90
Properties Non Polar aliphatic Non Polar aromathic Polar - Polar + Polar neutral
DQB1*06:02
DQB1*06:74
DQB1*06:01
DQB1*06:03
DQB1*06:08
DQB1*06:02
DQB1*06:74
DQB1*04:02
DQB1*04:01
-log10 (q-value)
aa position in peptide binding site
DQB1 locus

**Figure S7**

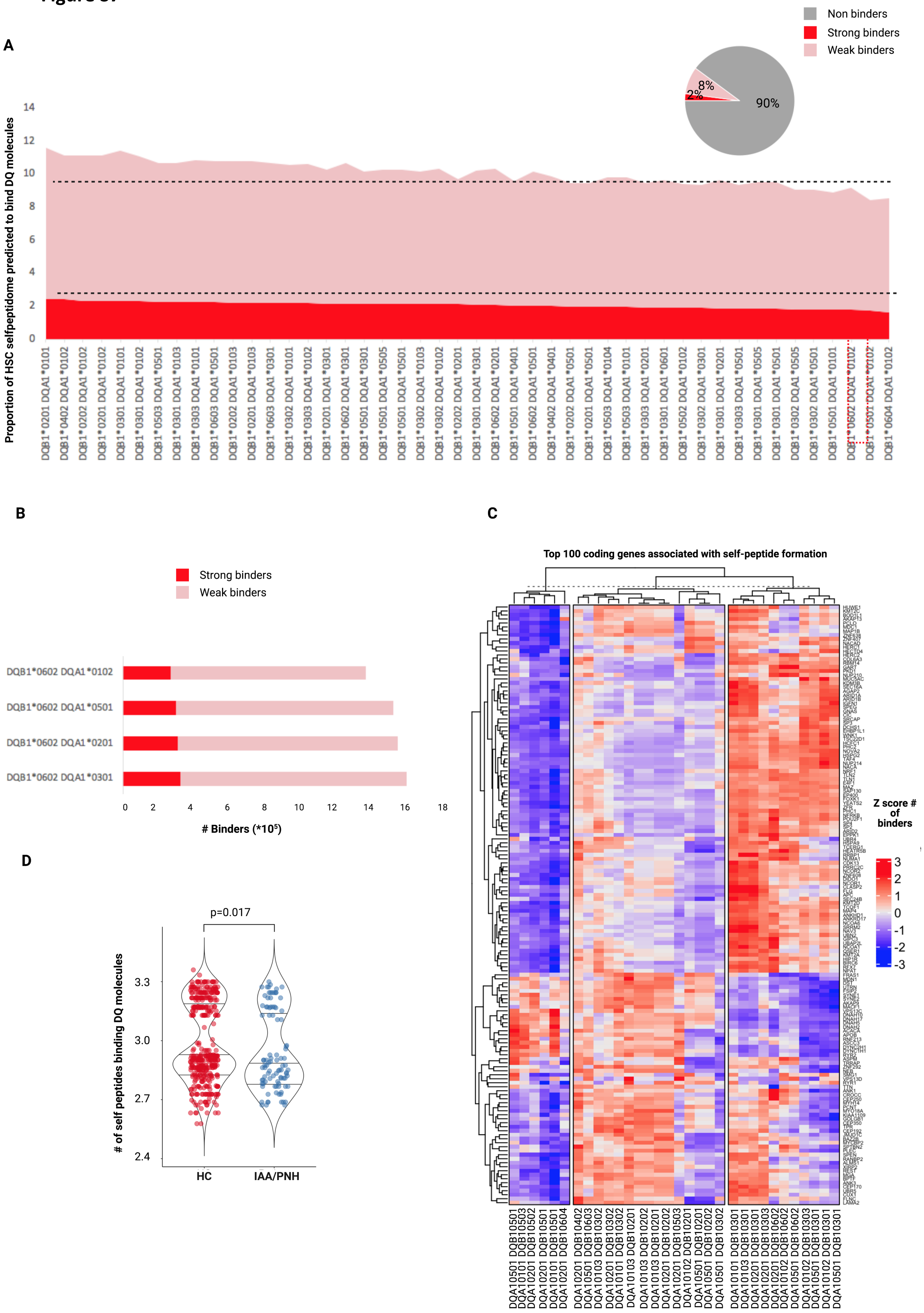
A
Non binders
Strong binders
Weak binders
90%
8%
2%
Proportion of HSC selfpeptidome predicted to bind DQ molecules
0
2
4
6
8
10
12
14
DQB1*0201 DQA1*0101
DQB1*0402 DQA1*0102
DQB1*0202 DQA1*0102
DQB1*0201 DQA1*0102
DQB1*0301 DQA1*0101
DQB1*0301 DQA1*0102
DQB1*0503 DQA1*0501
DQB1*0301 DQA1*0103
DQB1*0303 DQA1*0101
DQB1*0603 DQA1*0501
DQB1*0202 DQA1*0103
DQB1*0201 DQA1*0103
DQB1*0603 DQA1*0301
DQB1*0302 DQA1*0101
DQB1*0303 DQA1*0102
DQB1*0201 DQA1*0301
DQB1*0602 DQA1*0301
DQB1*0501 DQA1*0301
DQB1*0501 DQA1*0505
DQB1*0501 DQA1*0501
DQB1*0302 DQA1*0103
DQB1*0302 DQA1*0102
DQB1*0202 DQA1*0201
DQB1*0301 DQA1*0301
DQB1*0602 DQA1*0201
DQB1*0501 DQA1*0401
DQB1*0602 DQA1*0501
DQB1*0402 DQA1*0401
DQB1*0202 DQA1*0501
DQB1*0201 DQA1*0501
DQB1*0503 DQA1*0104
DQB1*0503 DQA1*0101
DQB1*0303 DQA1*0201
DQB1*0301 DQA1*0601
DQB1*0502 DQA1*0102
DQB1*0302 DQA1*0301
DQB1*0301 DQA1*0201
DQB1*0303 DQA1*0501
DQB1*0301 DQA1*0505
DQB1*0301 DQA1*0501
DQB1*0302 DQA1*0505
DQB1*0302 DQA1*0501
DQB1*0501 DQA1*0101
DQB1*0602 DQA1*0102
DQB1*0501 DQA1*0102
DQB1*0604 DQA1*0102
B
Strong binders
Weak binders
DQB1*0602 DQA1*0102
DQB1*0602 DQA1*0501
DQB1*0602 DQA1*0201
DQB1*0602 DQA1*0301
0
2
4
6
8
10
12
14
16
18
### Binders (*10⁵)
C
Top 100 coding genes associated with self-peptide formation
Z score # of binders
3
2
1
0
-1
-2
-3
D
p=0.017
### of self peptides binding DQ molecules
2.4
2.7
3.0
3.3
HC
IAA/PNH

**Figure S8**

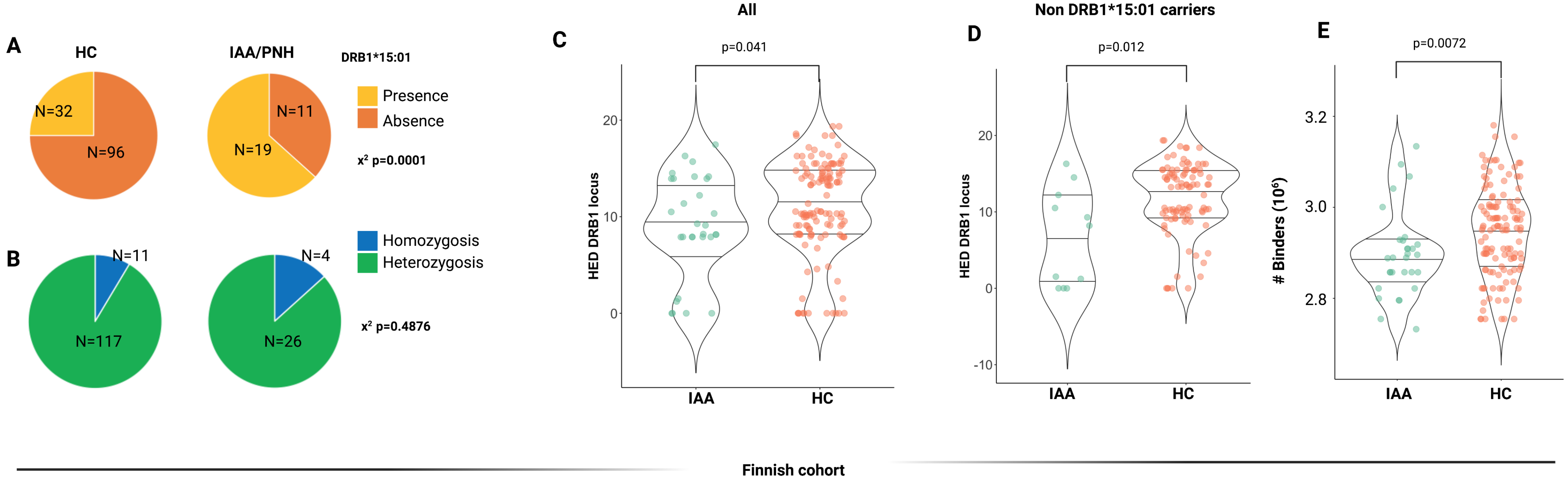
A
HC
IAA/PNH
DRB1*15:01
Presence
Absence
N=32
N=96
N=11
N=19
x² p=0.0001
B
Homozygosis
Heterozygosis
N=11
N=117
N=4
N=26
x² p=0.4876
All
C
p=0.041
HED DRB1 locus
IAA
HC
Non DRB1*15:01 carriers
D
p=0.012
HED DRB1 locus
IAA
HC
E
p=0.0072
### Binders (10⁶)
IAA
HC
Finnish cohort

#### Figure S9

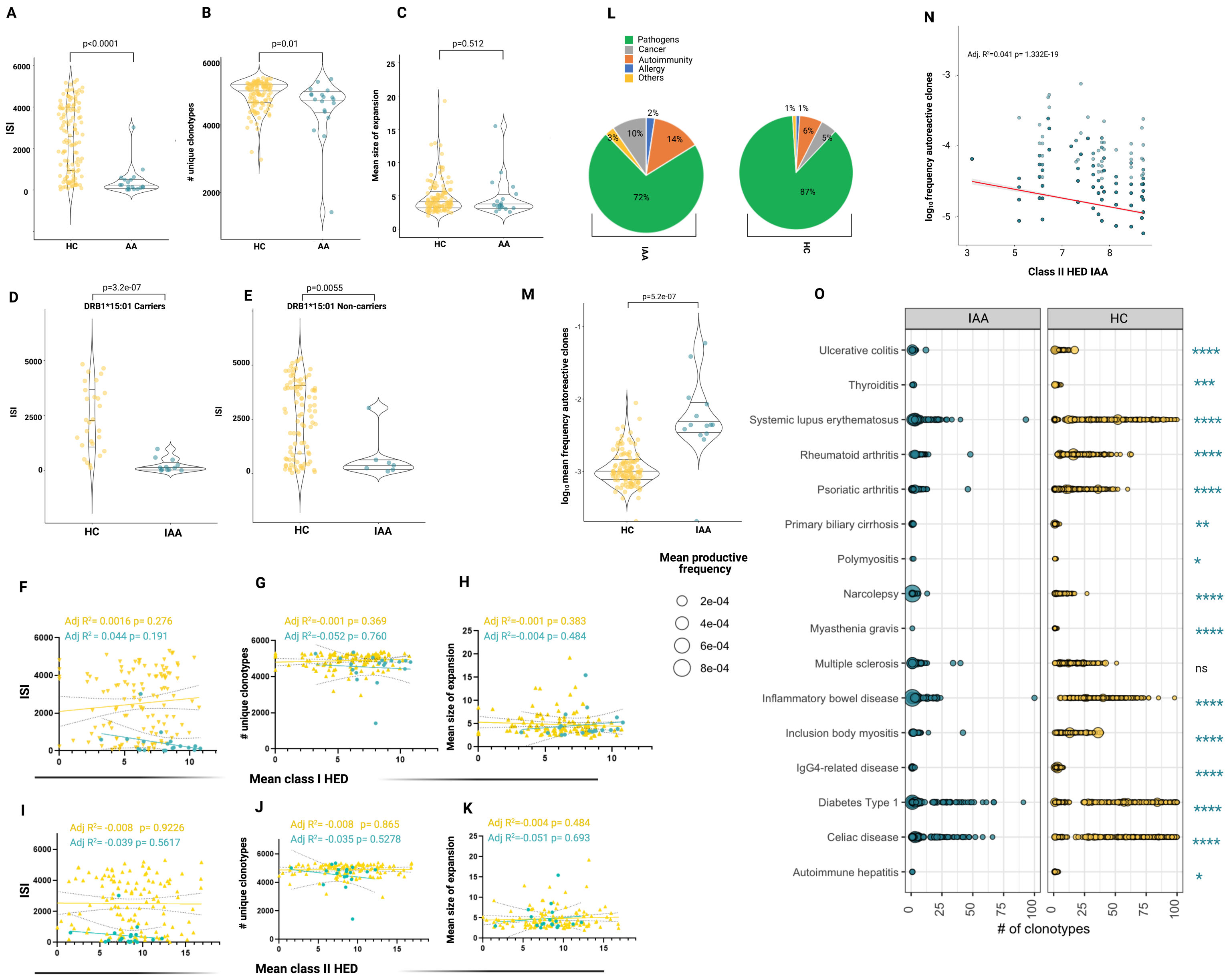

A
p<0.0001
ISI
HC
AA
B
p=0.01
### unique clonotypes
C
p=0.512
Mean size of expansion
L
Pathogens
Cancer
Autoimmunity
Allergy
Others
72%
14%
10%
3%
2%
87%
6%
5%
1% 1%
IAA
HC
N
Adj. R²=0.041 p= 1.332E-19
log₁₀ frequency autoreactive clones
Class II HED IAA
D
p=3.2e-07
DRB1*15:01 Carriers
E
p=0.0055
DRB1*15:01 Non-carriers
IAA
M
p=5.2e-07
log₁₀ mean frequency autoreactive clones
O
Ulcerative colitis
Thyroiditis
Systemic lupus erythematosus
Rheumatoid arthritis
Psoriatic arthritis
Primary biliary cirrhosis
Polymyositis
Narcolepsy
Myasthenia gravis
Multiple sclerosis
Inflammatory bowel disease
Inclusion body myositis
IgG4-related disease
Diabetes Type 1
Celiac disease
Autoimmune hepatitis
ns
### of clonotypes
Mean productive frequency
2e-04
4e-04
6e-04
8e-04
F
Adj R² = 0.0016 p= 0.276
Adj R² = 0.044 p= 0.191
G
Adj R²=-0.001 p= 0.369
Adj R²=-0.052 p= 0.760
H
Adj R²=-0.001 p= 0.383
Adj R²=-0.004 p= 0.484
Mean class I HED
I
Adj R²= -0.008 p= 0.9226
Adj R²= -0.039 p= 0.5617
J
Adj R²= -0.008 p= 0.865
Adj R²= -0.035 p= 0.5278
K
Adj R²=-0.004 p= 0.484
Adj R²=-0.051 p= 0.693
Mean class II HED

**Figure S10**

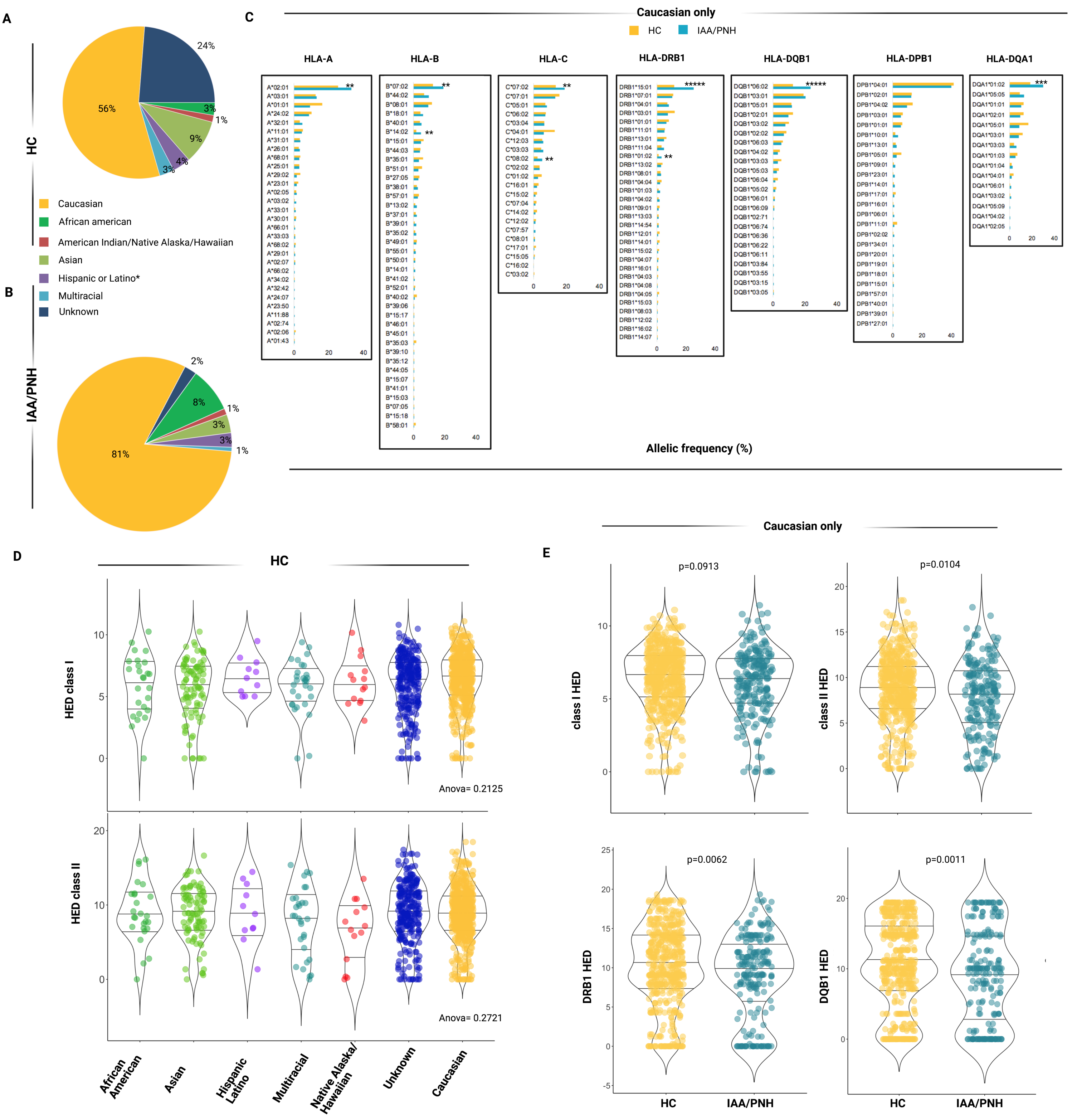
A
B
C
D
E
HC
IAA/PNH
Caucasian
African american
American Indian/Native Alaska/Hawaiian
Asian
Hispanic or Latino*
Multiracial
Unknown
Caucasian only
HLA-A
HLA-B
HLA-C
HLA-DRB1
HLA-DQB1
HLA-DPB1
HLA-DQA1
Allelic frequency (%)
HED class I
HED class II
Anova= 0.2125
Anova= 0.2721
African American
Asian
Hispanic Latino
Multiracial
Native Alaska/ Hawaiian
Unknown
Caucasian
class I HED
class II HED
DRB1 HED
DQB1 HED
p=0.0913
p=0.0104
p=0.0062
p=0.0011

**Figure S11**

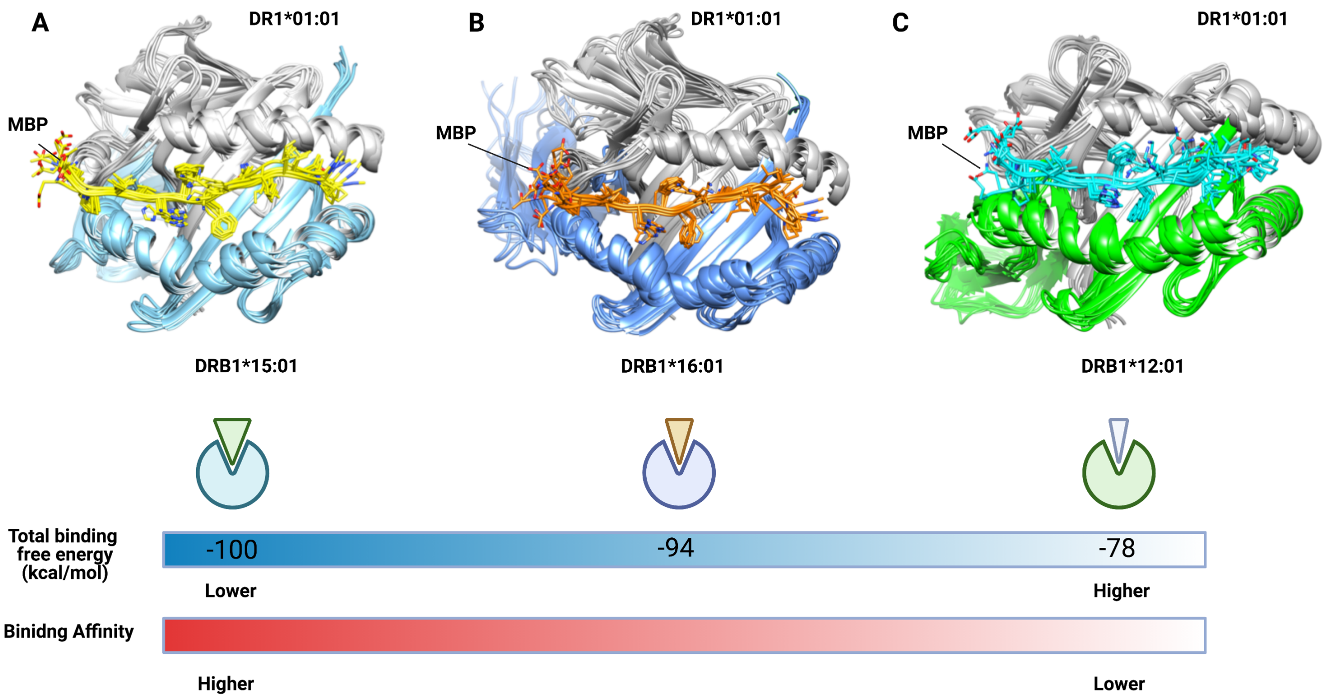
A
DR1*01:01
MBP
DRB1*15:01
B
DR1*01:01
MBP
DRB1*16:01
C
DR1*01:01
MBP
DRB1*12:01
Total binding free energy (kcal/mol)
-100
-94
-78
Lower
Higher
Binidng Affinity
Higher
Lower
